# Distinct modulatory effects of high-fiber and fermented-food diets on gut microbiota, immune function, transit time, and sleep quality in a citizen science randomized controlled trial

**DOI:** 10.1101/2025.08.05.25332853

**Authors:** Maartje van den Belt, Marieke van de Put, Ecem Yüksel, Donghyeok Lee, Fleur van Eeden, Jan Philip van Straaten, Nika Pintar, Bruno Balen, Mirna Andelic, Iris Rijnaarts, Isolde Besseling-van der Vaart, Marianne Rook, Douwe Molenaar, Evgeni Levin, Willem M. de Vos, Nicole J.W. de Wit, Remco Kort

## Abstract

A randomized, placebo-controlled citizen science-based dietary intervention was conducted among 147 healthy adults to evaluate the effects of 8-week high dietary fiber (HDF) and high fermented food (HFF) diets on gut microbiota, immune function, gut transit time, well-being, and sleep quality. The HDF group significantly increased fiber intake (Δ10.3 g/1000 kcal/day) following high dietary fiber recipes with addition of dried chicory root, while the HFF group increased fermented food consumption (+6.3 portions/day), including a fermentation-derived liquid supplement. At the 21-week follow-up, modest improvements in fiber and fermented intake were sustained, compared to baseline. Microbial diversity significantly increased within the HFF and control groups, especially in HFF participants over 50 (*p* = 0.04). Compared to CG, HFF showed no difference in microbial diversity, whereas the HDF group showed a significant decrease. The HDF intervention enhanced butyrogenic potential by increasing *Anaerostipes, Faecalibacterium*, and *Bifidobacterium* spp., and significantly reduced gastrointestinal transit time (*p* = 0.01). The intake of high fiber improved and sustained sleep quality (*p* = 0.03). The HFF intervention significantly increased blood immune markers including CD5, CD6 and CD8A (T-cell activation), IL-18R1 (inflammatory signaling) and SIRT2, a longevity-associated deacetylase (Q < 0.05), and induced a modest shift in the gut microbiota of participants over 50 years toward a composition characteristic of younger participants. These findings highlight distinct biological pathways through which dietary fibers and fermented foods modulate host physiology. This is the first randomized controlled nutritional intervention using a citizen science approach that demonstrates the feasibility and scientific value of engaging participants in healthier food choices.

**Graphical abstract:** 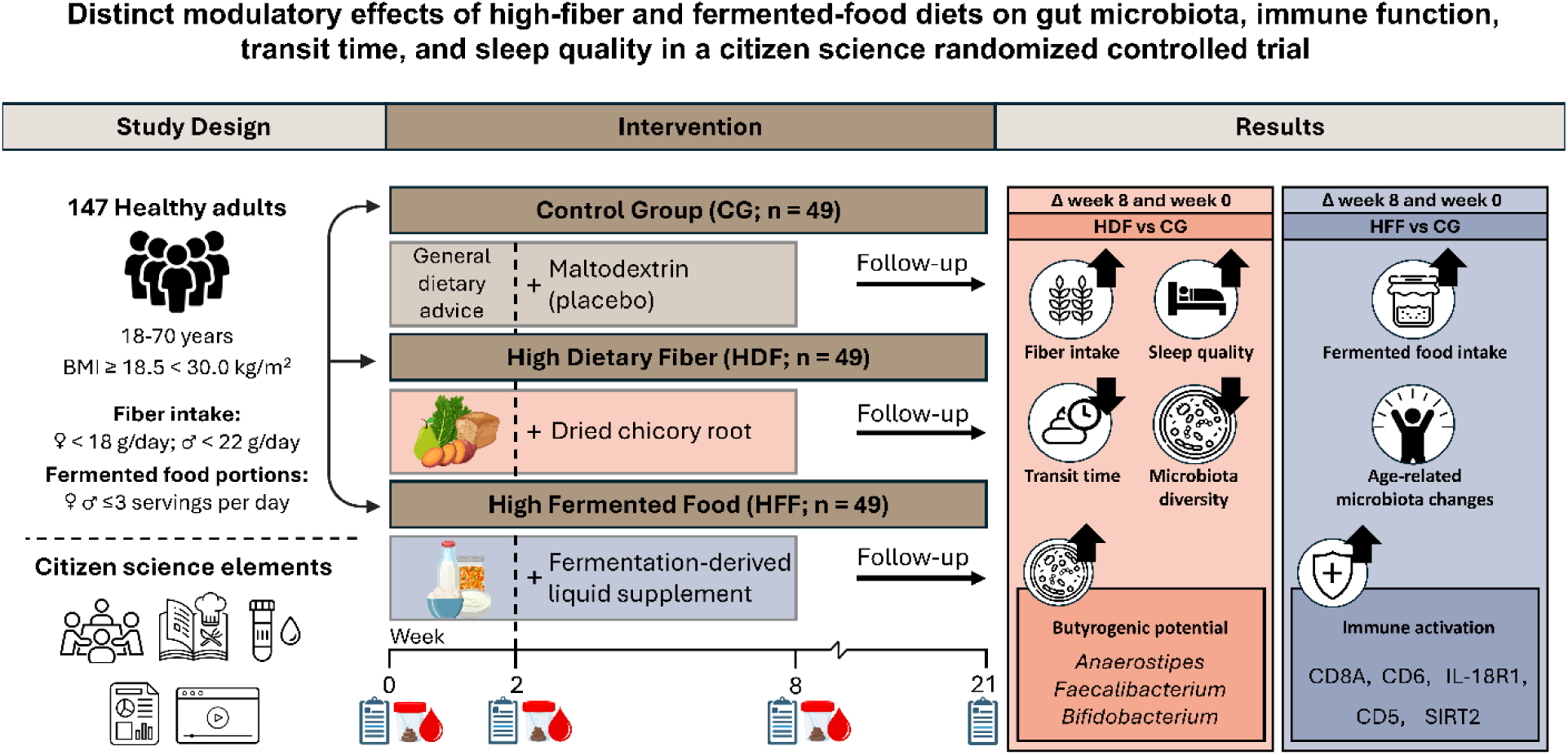

## 1. Introduction

With the global rise in diet-related chronic diseases, evidence-based dietary guidelines play a crucial role in promoting health (Armet et al., 2022). The human gut microbiota play an important role in both preventive health and disease (Joos et al., 2024; Ross et al., 2024). Dietary habits and lifestyle choices are important factors that shape the composition of the gut microbiota in adulthood (Rinninella et al., 2019). Moreover, gut transit time and stool consistency appear to be a major driver of gut microbiota variation, which is often overlooked in studies investigating the gut microbiota (Procházková et al., 2023; Vandeputte et al., 2016). There is a need for well-designed studies to better understand the relationship between diet, host health and the gut microbiota (Ross et al., 2024). There is no definition of a healthy microbiota yet, instead, various microbiota patterns may be associated with health and well-being (Joos et al., 2024; Shanahan et al., 2021; Van Hul et al., 2024).

A reduced microbial diversity is often linked to low-grade inflammation (Carding et al., 2015; Shivakoti et al., 2022; Wastyk et al., 2021; Zheng et al., 2020) and a higher risk of inflammation and frailty in elderly (Claesson et al., 2012). The gut microbiota and immune system are closely linked, with changes in the microbiota influencing metabolite production, intestinal tissue function, cellular activity and inflammatory responses (Clemente et al., 2018). In turn, the immune system also shapes microbiota composition by targeting and suppressing harmful microbes (Bosco & Noti, 2021). Hence, it is crucial to identify which food components and dietary patterns promote health via these mechanisms in order to develop practical dietary recommendations for health promotion and disease prevention (Ross et al., 2024).

Long term variations in dietary fiber intake have shown significant differences in gut microbiota diversity and composition (Makki et al., 2018). Therefore, dietary fiber is one of the most frequently studied for its impact on the gut microbiota (Wang et al., 2020). Most of these interventions include single, extracted dietary fibers, such as inulin or pectin (Yüksel et al., 2024). Whereas unprocessed fiber structures (intrinsic fibers) might enhance the gut microbiota and immune system more effectively due to their gradual release in the colon, as recently has been demonstrated for intrinsic chicory root fibers (Omary et al., 2025; Puhlmann & de Vos, 2022). Whole-diet approaches, such as the mediterranean diet or fiber-rich foods like whole grains, have shown beneficial effects on the gut microbiota composition and overall health (Armet et al., 2022). Fermentation of certain dietary fibers by the gut microbiota in the colon leads to the production of short-chain fatty acids (SCFA), which are involved in various human metabolic pathways and complex regulatory mechanisms to strengthen the intestinal barrier, impact the immune system, improve glucose and lipid metabolism, and support brain-gut axis communication (Kim, 2023; Roy et al., 2006; Tan et al., 2014; Yao et al., 2022).

Fermented foods represent another dietary component that recently attracted the attention of scientists for their potential impact on the gut microbiota (Shah et al., 2023). These foods are attributed with several potential benefits for cardiovascular, metabolic and immune health, largely due to the bioactive compounds and microbial metabolites produced during the fermentation process (Leeuwendaal et al., 2022). Furthermore, fermented foods that contain live micro-organisms can transiently introduce new microbes into the gut microbiota and serve as a significant dietary source of beneficial microbial species (Marco et al., 2017). Despite not being included in most national dietary guidelines, the incorporation of fermented foods could potentially restore crucial interactions between the gut microbiota and immune system, that have been diminished by modern lifestyles (Bell et al., 2017; Marco et al., 2020).

However, there is still limited knowledge about whole food interventions that effectively modulate the gut microbiota. Wastyk et al. previously investigated the effects of a high-fiber diet and a high-fermented-food diet on the gut microbiota and immune system in healthy adults (Wastyk et al., 2021). Over a ten-week intervention period, they observed that a diet rich in fermented foods consistently increased microbiota alpha diversity. In contrast, no significant changes in microbial diversity were detected in the high-fiber diet group. However, participants consuming a high-fiber diet exhibited an increase in microbiome-encoded glycan-degrading carbohydrate-active enzymes (CAZymes), suggesting enhanced carbohydrate metabolism. Furthermore, the fermented food intervention was associated with a reduction in systemic inflammation, as indicated by decreased levels of 19 cytokines in blood serum. In the high-fiber diet group, immunological responses varied based on baseline microbial diversity, revealing three distinct immunological trajectories. Although groups were randomized, the study lacked a non-intervention control group and had relatively small treatment arms (*n*=18 per group), limiting the generalizability of its findings. Larger randomized controlled trials are necessary to validate these results in other populations.

To effectively promote health through dietary recommendations, sustained long-term dietary modifications are essential. However, achieving such behavioral changes at the population level remains a significant challenge (Bowen, 1995; Kumanyika et al., 2000). Citizen science (CS) has the potential to engage, educate and empower study participants, fostering adherence to healthier dietary habits while raising public awareness of the impact of nutrition on the gut microbiota and its subsequent role in maintaining homeostasis and preventing disease (Den Broeder et al., 2016; Garcia et al., 2023; van de Put et al., 2024). Moreover, CS facilitates the scaling-up into public outreach programs, enabling broader dissemination of knowledge and strategies for improving gut health and the immune system at the community level. To explore this approach, we designed a randomized controlled trial (RCT) integrating CS elements (van de Put et al., 2024), comparing three dietary interventions: a high-dietary-fiber (HDF) diet with the addition of dried chicory root, a high-fermented-foods (HFF) diet with the addition of a fermentation-derived liquid supplement and a control group (CG) receiving maltodextrin. The Gut health Enhancement by Eating favorable Food (GEEF) trial will assess the effects of these diets on gut microbiota alpha-diversity and several other health related markers such as immune function, gut microbiota composition, stool pattern, sleep quality, and long-term dietary behavior changes.

## 2. Methods

### 2.1 Study design

We performed a 21-week, single blinded RCT consisting of three arms, in a parallel design. The study was conducted completely remote, between May 2023 and March 2024 in the Netherlands. The study protocol has been published previously (van de Put et al., 2024). In short, a high-dietary fiber intervention (HDF) and a high fermented foods intervention (HFF) were compared to a control group (CG). The study involved a 2-week ramp-up period followed by a 6-week intervention period, and a follow-up at week 21. Throughout the ramp-up and intervention period (8 weeks in total), participants in the HDF and HFF groups followed dietary guidelines based on recipe booklets, whereas the CG group only received general dietary advice by referring to specific websites. Additionally, all participants consumed a complementary study product during the 6-week intervention period. Several elements of citizen science were adopted in the study design, to empower participants with knowledge and to motivate them towards sustaining dietary changes.

Study assessments were performed during the following four timepoints: prior to the study start (week 0, baseline), at week 2 (end ramp-up), at week 8 (end intervention) and at week 21 (follow-up). Baseline, week 2 and week 8 measurements included the collection of fecal samples, dried blood spot samples, stool smear images, assessment of gut transit time and completion of questionnaires on well-being, sleep, perception and awareness of food choices, and quality of life. Additionally, participants completed a 3-day food diary using the Traqq application on their smartphones (Lucassen et al., 2021). Daily questionnaires on their mobile phone assessed stool pattern, gastrointestinal complaints and compliance to the dietary guidelines and study product during the 8-week period. The follow-up measurement consisted of the questionnaires and the 3-day food diary. Figure 1 shows a schematic overview of the study design.

**Figure 1.**
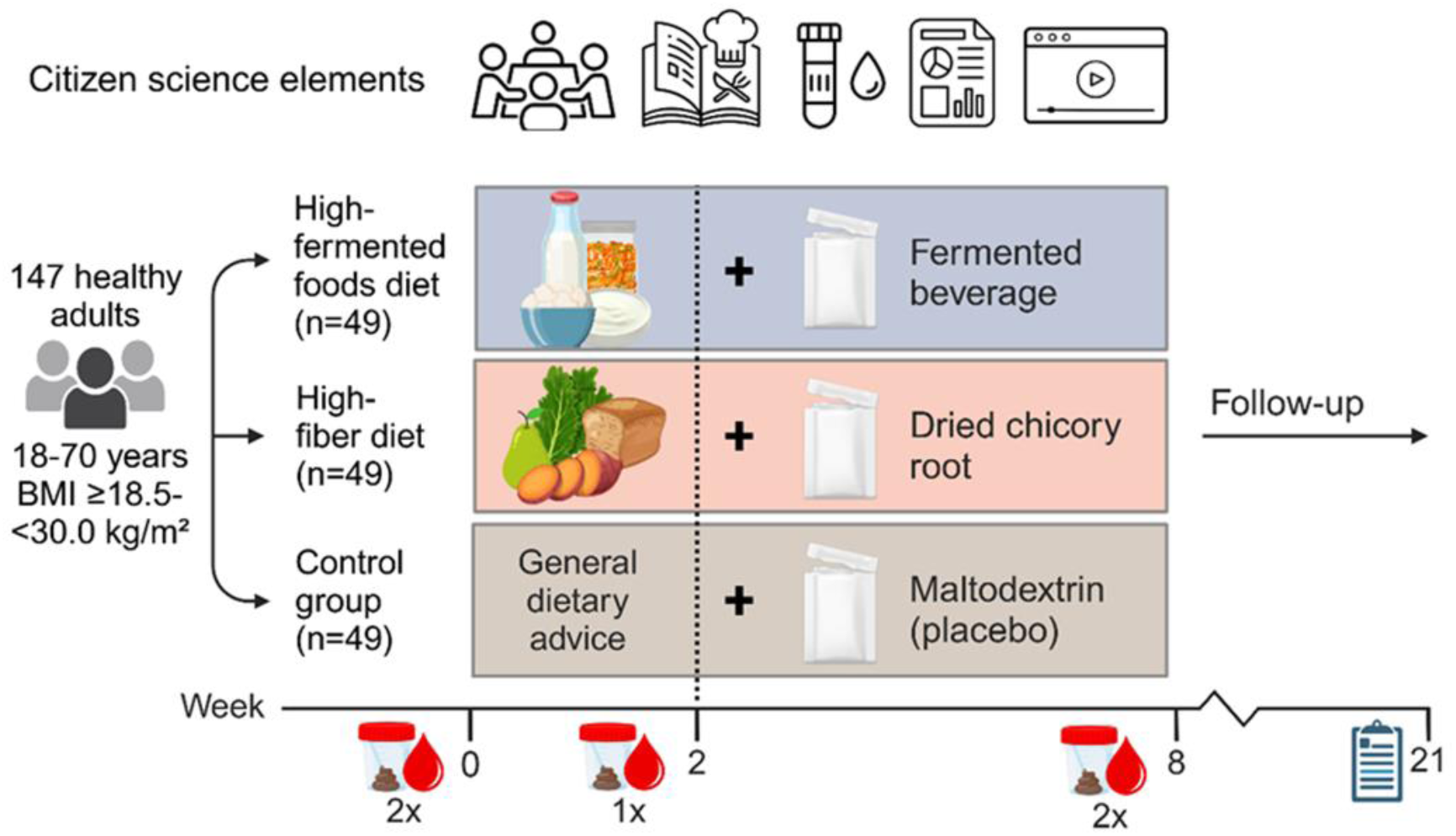
Schematic overview of the study design of the GEEF trial including healthy adults. A total of 147 healthy adults were randomized to an 8-week high-dietary fiber diet, high-fermented foods diet, or control group. Measurements were performed during baseline (week 0), end ramp-up (week 2), end intervention (week 8) and follow-up (week 21). Citizen science elements included education, a tailored dietary intervention, at-home sampling by participants, provision of personal gut microbiota reports, and participant-created vlogs for dissemination of findings. Figure copied from our study protocol paper by van de Put et al. (2024).

Before the start of the study, participants received the recipe booklets and all collection materials as described in our study design paper (van de Put et al., 2024). Online separate start-up meetings for each treatment group were organized to inform participants on all study procedures and measurements. Additionally, study websites were created for each treatment group where participants could find written and video recorded information on study measurements and the importance of the gut microbiota. To facilitate working with this large sample size, two starting groups were used. The first group started in June 2023 and completed the study in February 2024, the second group started in September 2023 and completed the study in March 2024.

### 2.2 Study participants, randomization and blinding

Based on the sample size calculation, which is explained in our study design paper (van de Put et al., 2024), we aimed to include 147 study participants. Participants aged ≥18 and ≤70 years with a BMI ≥18,5 and <30 kg/m^2^ were considered. Main exclusion criteria were having a medical condition influencing study outcomes, antibiotic use within three months prior to study start or relatively high dietary fiber intake (≥18 g/day for women or ≥22 g/day for men, screened by using the FiberScreen questionnaire (Rijnaarts, de Roos, Zoetendal, et al., 2021), or fermented food intake (>3 servings per day). Stratified permuted block randomization was used to equally divide the eligible participants over the treatment groups, and to balance for sex, age and BMI. Participants in the HDF and HFF groups were informed on their specific dietary advice, however they were unaware of how their treatment differed from the other groups. The CG was blinded until the end of the 8-week intervention. All treatment groups were blinded to the content of the study sachets, holding the complementary study product per treatment. After follow-up, group labels were re-coded by an independent person, enabling blinded data-analysis. For all inclusion and exclusion criteria and details on recruitment, randomization and blinding we refer to the study design paper (van de Put et al., 2024).

### 2.3 Dietary interventions

Briefly, the HDF group was instructed to increase dietary fiber intake by daily incorporation of two high-fiber snacks and following two high-fiber meal recipes from the recipe booklet. We aimed to increase dietary fiber intake, using the recipe booklets, by on average 24 g fiber/day. Participants in the HFF group were instructed to add three extra servings of fermented foods to their daily diet. The participants could select their servings from a product list, containing products with live micro-organisms and their corresponding serving sizes, which could be either store-bought or homemade. All three treatment groups received general information about diet in relation to the gut microbiota by referring them to the websites of the Dutch Digestive Health Fund and The Netherlands Nutrition Centre. In the CG group this sole and general dietary information is unlikely resulting in substantial dietary modifications (Rijnaarts, de Roos, Wang, et al., 2021). Besides the dietary guidelines, participants were instructed to consume three study sachets per day during the 6-week intervention period. The sachets for the HDF group consisted of 3.3 grams dried chicory root (WholeFiber^TM^, WholeFiber Holding BV, The Netherlands) (Puhlmann et al., 2022), resulting in an additional ∼7.9 g fiber/day. When combined with the recipe booklets recommendations, the estimated total fiber intake during the intervention ranged from 27 to 37 g fiber/day. Dried chicory root consists of a mixture of inulin, pectin and (hemi-)cellulose still in the intrinsic plant cell wall and has been shown to have an effect on the gut microbiota and SCFA production (Puhlmann et al., 2022; Puhlmann & de Vos, 2020).

The HFF group received sachets containing 19 milliliters of a fermentation-derived liquid supplement Agebiotics^TM^ (Cidrani d.o.o., Croatia/ Ani Biome, Inc., USA), with the instruction to dilute this in a glass of water before consumption. See Table S1 and S2 for compositional details of the Agebiotics^TM^ product. Daily consumption of these three sachets facilitated the recommended consumption of six additional portions of fermented foods per day, as shown before to significantly increase alpha-diversity (Wastyk et al., 2021). The CG group received sachets containing 3.3 grams maltodextrin (Roquette Freres SA, France), providing a total daily intake of 9.9 g / day. More detailed nutritional values of the study products are reported in the protocol paper (van de Put et al., 2024). After the 8-week intervention, the study product was no longer administered, and the dietary guidelines (both the HDF and HFF recipe booklets) were made available to all participants without any obligations. At follow-up (week 21), dietary intake was assessed again to determine long-term changes in eating behavior. To evaluate effectiveness of the dietary intervention (wk0-8) and determine sustainability of dietary changes (wk21), dietary intake was assessed using the Traqq app on week 0, 2, 8 and 21.

### 2.4 Primary and secondary outcomes

The primary outcome measure was the change in gut microbiota alpha diversity from baseline following the 8-week dietary intervention, determined from fecal samples assessed using V3-V4 16S rRNA gene amplicon sequencing. Microbiota data analyses were carried out using QIIME2 version 2023.2.0 (Bolyen et al., 2019). QIIME workflows were run under Snakemake version 7.32.3. (Köster & Rahmann, 2012; Mölder et al., 2021). Modified workflows, scripts and environments were used from Snaq (Mohsen et al., 2022). The read statistics were compiled using MultiQC (Ewels et al., 2016). Low quality bases were removed using bbduk (BBTools - DOE Joint Genome Institute, 2023). Two thresholds (trimq=16 and trimq=20) were compared by their effect on the final number of non-chimeric sequences. A threshold of 20 yields slightly better results, therefore, quality threshold trimq=20 was used. Primer sequences were removed using fastp, 17 bases in the forward sequence and 21 in the reverse sequence (Chen et al., 2018). Data was denoised and unique amplicon sequence variants (ASVs) were identified using DADA2, version 1.26 (Callahan et al., 2016). Alpha diversity was calculated after rarefying the data to 10,000 reads per sample, retaining 513 out of 567 samples. For relative abundance analyses, all samples were included; however, amplicon sequence variants (ASVs) with over 80% sparsity were excluded to minimize the influence of potential sequencing artifacts (Nearing et al., 2021).

Phylogenetic positions were assigned to ASVs using the Silva V34 classification (Supplemental excel file S1). To obtain species-level information, included in the footnotes of the figures, ASVs were aligned against the NCBI Microbial Nucleotide Basic Local Alignment Search Tool with default parameters (Altschul et al., 1990). Taxonomic assignment was determined based on the top match with a minimum sequence identity threshold of 98.65 % for species-level identification (Kim et al., 2014). In cases of ambiguous matches, species classification was not assigned. Alpha diversity measures included Shannon diversity, Chao1, the abundance-based coverage estimators index (ACE), Fisher’s alpha and the number of observed ASVs. Very high correlation coefficients (> 0.99) were found for ACE, Fishers’ alpha, Chao and Observed ASVs. Therefore, in further analyses Shannon diversity and number of observed ASVs were used as alpha diversity measures.

A detailed description of the secondary outcomes has been reported previously in our study design paper (van de Put et al., 2024). In summary, these outcomes included 92 inflammatory protein biomarkers in dried blood spots, gut transit time, dietary intake, gut microbiota composition*, Prevotella* to *Bacteroides* ratio via the stool smear image tool, well-being, sleep quality, digestion-associated quality of life, perception and awareness of food choices, stool consistency, stool frequency, and gastrointestinal complaints (bloating, flatulence, and abdominal cramping). An overview of all study outcomes and timepoints can be found in Table S3.

### 2.5 Statistical analysis

Statistical analysis and visualizations were performed using R statistics (Version 4.4.1) and Python (Version 3.11.9). *p*-values of <0.05 were considered statistically significant. For the primary outcome (alpha diversity) and secondary outcomes linear mixed models (LMM) were used, unless specified otherwise below. Data were fitted using the nlme package to analyze trends in gut microbiota diversity, with as fixed effects treatment group [HDF/HFF/CG], time point [week 0/week 8], and as interaction treatment*time. Slope and offset were modeled as subject-dependent random effects, nested within the treatment groups. Potential associations between personal characteristics (sex, BMI and age) and baseline alpha-diversity scores were investigated visually (for sex) and using linear regression models (for BMI and age). Since there was a positive trend between age and alpha diversity *(p*=0.06), further exploratory analysis was performed modeling age (≤50 and >50) as a triple interaction effect with treatment and time. Samples with relatively low read counts (<10,000 reads) were excluded from the bacterial diversity analyses as they have a higher likelihood of missing low-abundance microbial species, leading to an underestimation of species richness and diversity indices.

For microbiota analysis (at ASV and genus-level taxon) and immune markers, an adjusted *p*-value, corrected for multiple testing using the Benjamini-Hochberg false discovery rate (Q-value), was reported. Within group differences were tested using Wilcoxon signed rank tests on paired samples. For between-group comparisons (HDF and HFF compared to CG), the delta for each marker was calculated as the difference between week 8 and week 0 for each paired sample, followed by analysis using the Mann–Whitney U test. Finally, volcano plots were generated for the immune markers to visualize both statistical significance and effect size. For the analysis of the immune markers, only the 69 protein markers detected in more than 75% of samples, out of a total of 92 proteins, were included in the analysis. The results were further supported by a heatmap based on the delta values of the resulting significant markers, to indicate group-level differences. For microbiota analysis, a heatmap was generated by aggregating ASV counts by genus and log-transforming the data, including only taxonomic assignments with a confidence score >0.9. Taxa with at least 50 sequence counts in any single individual at any time point were included. Taxonomic units were ordered by decreasing relative abundance, summed over all samples (Supplemental file S3 heatmap).

Other secondary outcome parameters (stool smear images, transit time, dietary intake, sleep quality, digestive associated quality of life, well-being and the different scales for perception and awareness) were analyzed using LMM with restricted maximum likelihood estimation using the lmer function (“lme4” package in R). Treatment group [HDF/HFF/CG], and time point [week 0/week 2/week 8/week 21] were included in the model as main effects and as interaction effect (treatment*time), and participant was included as random effect. Week 0 was included in the model as the reference. Additionally, a model was run with week 2 as the reference, to distinguish between the effects of the ramp-up and intervention periods. One subject was excluded from the transit time analysis at baseline due to an extreme outlier, defined as a value exceeding the mean by more than three standard deviations (mean + 3 SD) at one of the three measurement time points.

To test if participants were able to sustain changes after the intervention period, outcome measures at week 21 were compared to week 0, within-groups, using a paired t-test.

The outcome measures for gastrointestinal complaints, stool frequency and stool consistency were collected on a daily base during the ramp-up and intervention period, resulting in many repeated measurements within individuals. For these outcome measures LMM was used with treatment group and time (baseline [day 0 - 2]/ramp-up [day 3-14]/intervention [day 15-56]) included in the model as main effect and interaction effect, and the subject as random effect.

#### 2.5.1 Machine learning

To assess changes in microbiome composition following the intervention, a multivariate classifier model was developed using delta values (week 8 minus week 0) to distinguish between the CG and the treatment groups. Model performance was evaluated using the Area Under the Receiver Operating Characteristic curve (AUC). The extremely randomized trees algorithm (Geurts et al., 2006) was employed, with the following pipeline: the dataset was randomly split 50 times into 70% training and 30% test sets using stratified shuffle splits to ensure balanced class distributions. For each split, the 100 features with the highest variance were selected from the training set to reduce data dimensionality and capture markers with large effect sizes. Hyperparameters (maximum depth: 2–4; minimum samples per split: 3–5; minimum samples per leaf: 3– 5) were optimized using 5-fold stratified inner cross-validation to maximize the AUC. To evaluate the statistical significance of model performance, a permutation test (Ojala & Garriga, 2009) was conducted by shuffling the labels 50 times, each time repeating the same model pipeline. The AUC was calculated for each permuted test set, and the p-value was computed as the proportion of permuted AUC scores meeting or exceeding the mean value of the original AUC scores. Finally, permutation importance (Altmann et al., 2010) was calculated on the test set with 100 permutations to identify key microbiome markers driving group differences. Based on the delta values of the top 10 ASVs resulting from this model, heatmaps were created.

For within-group analysis, the same procedure was applied based on the paired data, but a multiple-participants-out approach was used, ensuring that all samples from the same participant are assigned to either the training or test set simultaneously, as samples from the same participant are not independent of each other. The machine learning pipeline was implemented in Python (v3.11.9) using the scikit-learn (v1.5.1) package (Fabian Pedregosa et al., 2011).

#### 2.5.2 Exploratory analyses

Exploratory analyses were conducted to investigate differences in microbial composition in relation to gut transit time and age. The age-related analysis was included due to a positive association between age and the primary outcome, alpha diversity. Gut transit time was further explored given its established role as an important covariate contributing to the interindividual variation in gut microbiota composition (Procházková et al., 2023). Participants were stratified into long (top 30%, >32.93 hours, week 0,102 samples) and short (bottom 30%, <23.66 hours, week 0, 102 samples) transit time groups, as well as into younger (≤50 years, week 0, 63 participants) and older (>50 years, week 0, 50 participants) age groups across all study arms. These analyses were complemented by between-group comparisons (HDF vs CG, HFF vs CG), using delta values (week 8 minus week 0) and were statistically evaluated with the Mann–Whitney U test. Q-values were calculated using the Benjamini–Hochberg FDR method. For the age-related analysis, baseline differences in microbiota composition between participants younger and older than 50 years were first assessed across the three study groups. Subsequently, the 10 most important microbial markers distinguishing younger from older adults were identified and incorporated into a principal component analysis (PCA) to assess the impact of the dietary interventions on these ten age-associated microbial features.

#### 2.5.3 Population analysis

The intention to treat (ITT) population was defined as all participants that were randomized and received a treatment. The per protocol (PP) population was defined as those participants who completed the 8-week intervention without major protocol deviations; completed at least 75% of daily questionnaires, and also sufficiently complied to the intake of study sachets and dietary guidelines (intervention groups: ≥80% intake of study sachets and ≥80% adherence to dietary guidelines; CG: ≥80% intake of study sachets). The reported results are based on the PP analysis. The PP population for each outcome, along with the reasons for exclusion of participants from the PP population, is reported in Table S4.

## 3 Results

### 3.1 High adherence facilitated by citizen science design

A total of 147 eligible participants were enrolled in the study, 146 of whom received the allocated intervention (ITT population), and 133 completed the protocol without major deviations (PP population, Figure 2). Participants were geographically distributed across the Netherlands, covering nearly the entire country (Figure S1). Baseline characteristics were comparable across the three study arms (Table 1). The majority of participants were female (71–75% per group), with a mean age of 44.5 years and an average BMI of 24.1 kg/m². Baseline daily dietary fiber intake ranged from 20.3 to 22.1 g/day across groups, while fermented food consumption was uniformly low (0.6– 0.8 portions/day). Body weight remained stable throughout the intervention in all groups (Figure S2), indicating that any observed outcomes were unlikely to be confounded by weight change and could be attributed to the dietary interventions.

**Figure 2.**
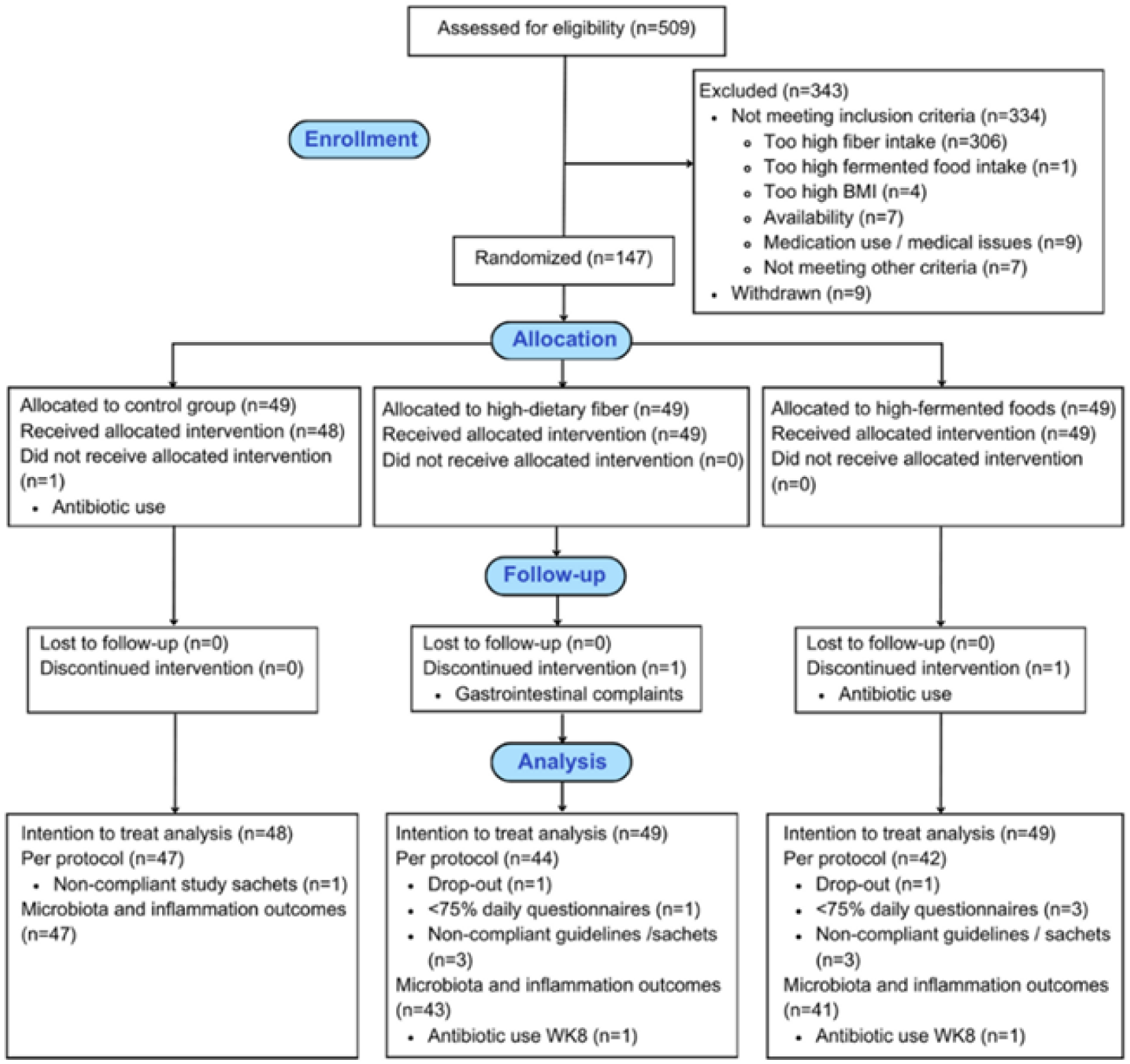
CONSORT flow diagram of study participants in the GEEF trial. Figure copied from the study protocol paper by van de Put et al. (2024).

**Table 1.** Baseline characteristics of the study participants in the GEEF trial including healthy adults aged 19-69 years, for the ITT population (*n*=146).

|  | **CG (n=48)** | **HDF (n=49)** | **HFF (n=49)** |
| --- | --- | --- | --- |
| Sex |  |  |  |
| Female, *n* (%) | 36 (75.0) | 36 (73.5) | 35 (71.4) |
| Male, *n* (%) | 12 (25.0) | 13 (26.5) | 14 (28.6) |
| Age (years), mean (± SD) | 45 (14.6) | 43 (14.0) | 46 (13.1) |
| BMI (kg/m<sup>2</sup>), mean (± SD) | 24.0 (2.3) | 23.8 (2.9) | 24.3 (2.6) |
| Fiber intake (g/day), mean (± SD) | 22.1 (10.0) | 20.3 (7.9) | 20.6 (9.6) |
| Fermented food intake (portions/day), mean (± SD) | 0.6 (0.9) | 0.6 (0.8) | 0.8 (1.2) |

Adherence to both the dietary guidelines and study product consumption was consistently high across all groups. Only 12 participants (8.2%) reported mild gastrointestinal complaints in relation to the intervention (*e.g.*, transient bloating or discomfort, HDF: n=1, HFF: n=7, CG: n=4). All complaints were short-lived and did not require any medical intervention (Table S5; Figure S3). The high adherence observed may be attributed to the citizen science approach of the study, which actively engaged participants and encouraged sustained dietary modifications. Additional details on recruitment, study design, and participant compliance are provided in our protocol publication (van de Put et al., 2024).

### 3.2 Dietary intervention compliance and behavior change

In the high-dietary fiber (HDF) group, baseline fiber intake was on average 20.4 g/day (11.3 g/1000 kcal). This significantly increased over the first two weeks by 4.8 g/1000 kcal, compared to the CG (*p*<0.001, Table S6, Figure S4). During the subsequent 6-week intervention period, which included both dietary advice and consumption of the dried chicory root study product, fiber intake in the HDF group increased by 10.3 g/1000 kcal relative to baseline (*p*<0.001, Figure 3). This resulted in an average dietary fiber intake of 37.8 g/day, or 21.6 g/1000 kcal, during the last six weeks of the intervention. Throughout the intervention, the primary sources of fiber in the HDF group included grains (9.0 g/day), vegetables (5.6 g/day), and fruit (3.6 g/day) (Figure S5). In contrast, fiber intake remained stable in the HFF group and showed a slight, non-significant decline in the CG (*p*=0.086). At follow-up, both the HDF and CG groups had significantly lower energy intake compared to baseline (*p*=0.005 and *p*=0.015, respectively, Table S6). After adjusting for energy intake, dietary fiber intake in the HDF group remained significantly higher compared to baseline (Δ=2.2 g/1000 kcal, *p*=0.004, Figure S5), with continued increased intake of fruits, grains, and nuts (Figure S6). No significant change in dietary fiber intake was observed in the CG (Δ=0.36 g/1000 kcal, *p*=0.437) or HFF group (Δ=0.07 g/1000 kcal, *p*=0.868) at follow-up.

**Figure 3.**
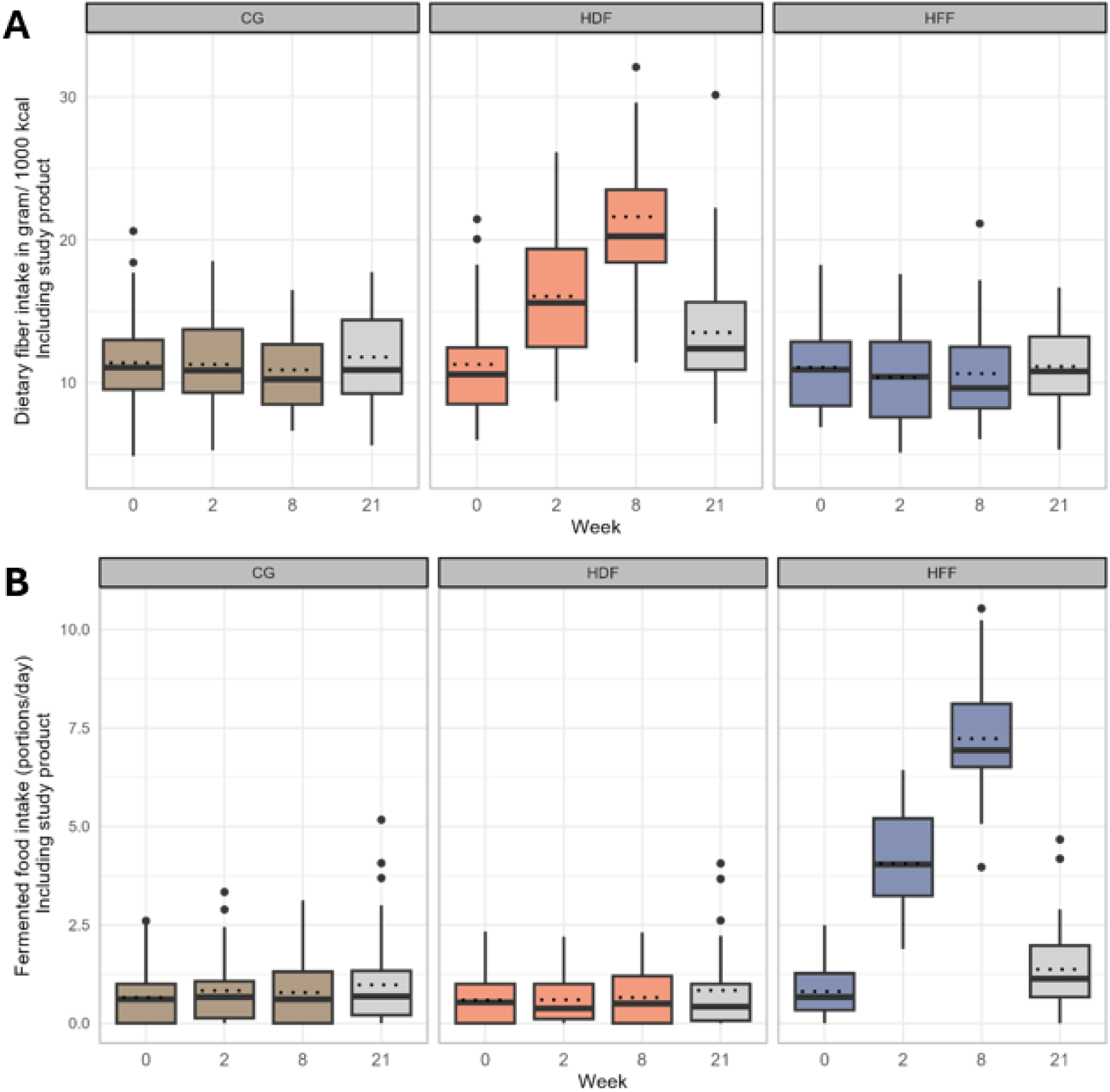
Dietary fiber intake (grams/day) and fermented food intake (portions/day) including the intake of the study products. Dietary intake was measured with 3-day food diaries using the Traqq application at baseline (week 0), end of ramp-up (week 2), end of intervention (week 8) and end of follow-up (week 21) for the per protocol population. Thick line shows the median and dotted line the mean value. Abbreviations: CG, control group; HDF, high-dietary fiber; HFF, high-fermented food; PP, per protocol.

Participants in the HFF group significantly increased their fermented food intake by 3.2 portions per day over the first two weeks compared to the CG (*p*<0.001, Figure S4). Over the subsequent 6-week intervention period, fermented food intake increased further, with participants consuming 6.3 additional portions per day relative to baseline and the change within the CG (*p*<0.001, Figure 3). This resulted in an average intake of 7.2 portions/day during the last six weeks of the intervention. The most commonly consumed fermented foods in the HFF group were fermented dairy (2.1 portions/day) and fermented drinks (1.1 portions/day) (Figure S7). Fermented food intake remained stable in both the HDF and CG groups throughout the intervention. At follow-up, fermented food intake remained significantly higher compared to baseline in the HFF group (Δ=0.56 portions/day, *p*=0.002). This increase was primarily driven by sustained consumption of fermented dairy (Figure S7). No significant changes in fermented food intake were observed in the CG (Δ=0.32 portions/day, *p*=0.051) or HDF group (Δ=0.25 portions/day, *p*=0.102) at follow-up. A summary of the dietary intake per treatment group is provided in Table S6.

### 3.3 Modulation of gut microbiota and compositional shifts

Gut microbiota alpha diversity, measured by Shannon diversity, is shown in Figure 4 for each treatment group across the 8-week intervention. A significant within-group increase in Shannon diversity was observed in both the control group (CG; *p* = 0.009) and the high-fermented food (HFF) group (*p* = 0.03), whereas diversity did not change within the high-dietary fiber (HDF) group. However, linear mixed model analysis indicated a modest but statistically significant decline in Shannon diversity in the HDF group in relation to the change within the CG over time (Table 2; *p* = 0.03).

**Figure 4.**
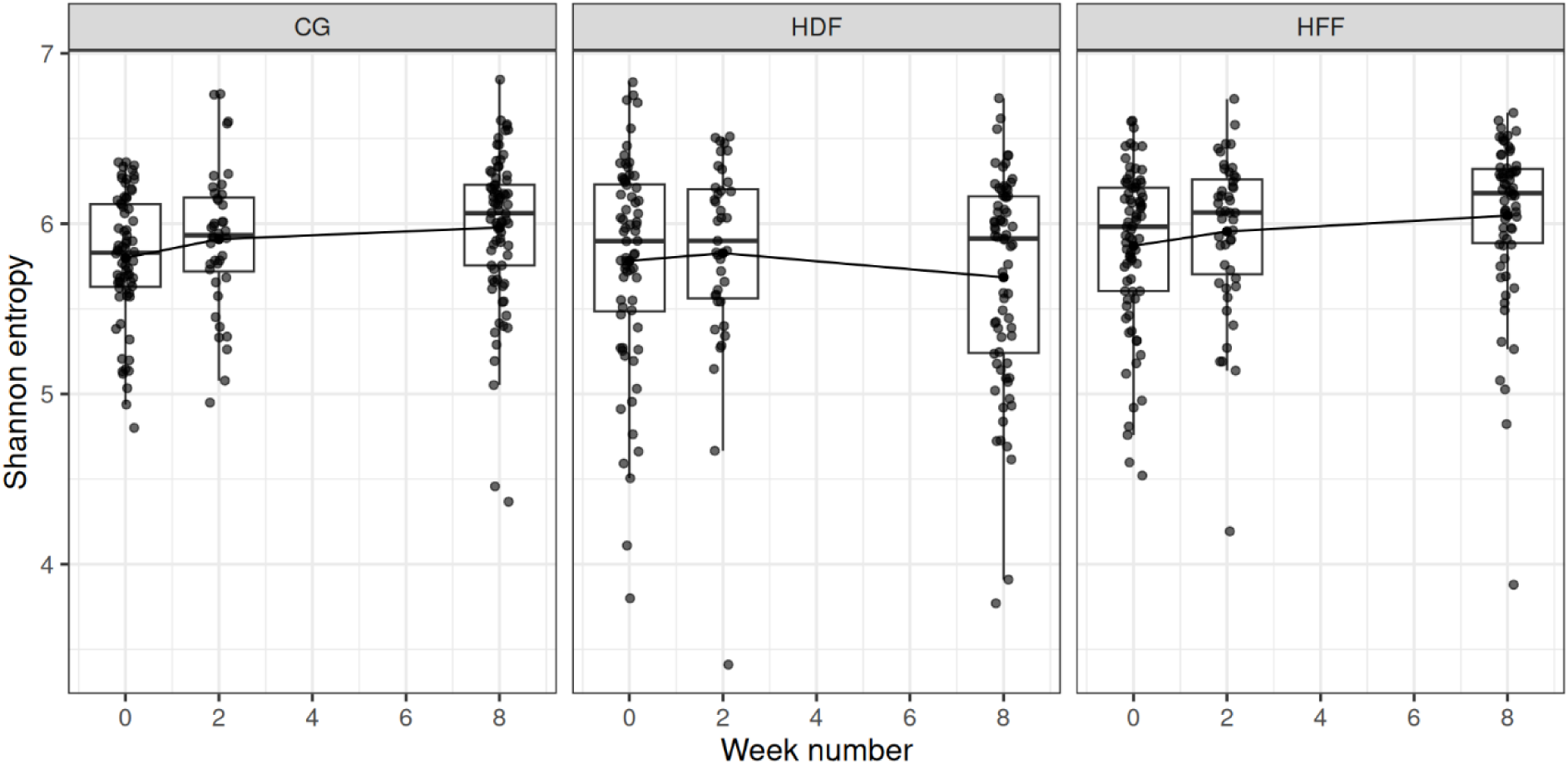
Development of alpha diversity, determined by Shannon diversity over time in the CG, HDF group, and HFF group of the GEEF trial for the PP population. Abbreviations: CG, control group; HDF, high-dietary fiber; HFF, high-fermented foods; PP, per protocol. All samples with less than 10,000 reads were excluded from this analysis.

**Table 2.** Primary and secondary outcomes related to gut microbiota and stool pattern, comparing the CG, HDF group and HFF group of the GEEF trial, for the PP population.

| Week | CG |  |  | HDF |  |  | HFF |  |  | Change between HDF and CG<br>(week 0-8) |  | Change between HFF and CG<br>(week 0-8) |  |
| --- | --- | --- | --- | --- | --- | --- | --- | --- | --- | --- | --- | --- | --- |
|  | 0 | 2 | 8 | 0 | 2 | 8 | 0 | 2 | 8 | Treatment effect<br>(95% CI) | P-value | Treatment effect<br>(95% CI) | P-value |
| Observed number of ASVs, mean (SD) | 117 (28) | 126 (34) | 131 (25)* | 125 (38) | 121 (31) | 119 (30) | 124 (35) | 127 (32) | 135 (25) | -2.210 (-4.283 to -0.138) | 0.037 | -0.170 (-2.241 to 1.901) | 0.87 |
| Alpha diversity (Shannon index), mean (SD) | 5.80 (0.38) | 5.91 (0.42) | 5.98 (0.44)* | 5.78 (0.62) | 5.83 (0.59) | 5.69 (0.62) | 5.87 (0.47) | 5.95 (0.46) | 6.05 (0.48)** | -0.039 (-0.073 to -0.005) | 0.025 | -0.005 (-0.039 to 0.029) | 0.77 |
| Ratio of *Prevotella/Bacteroides* (SD) | 0.456 (0.261) | 0.446 (0.189) | 0.674 (0.386) | 0.688 (0.436) | 0.481 (0.518) | 1.03 (0.501) | 0.676 (0.277) | 0.660 (0.551) | 0.450 (0.535) | 0.013 (-0.045 to 0.071) | 0.66 | -0.034 (-0.097 to 0.030) | 0.30 |
| Alpha diversity+ (SD) | 5.75 (0.39) | 5.85 (0.41) | 5.83 (0.45) | 5.83 (0.51) | 5.83 (0.52) | 5.66 (0.62) | 5.83 (0.50) | 5.92 (0.42) | 5.90 (0.50) | -0.030 (-0.057 to -0.002) | 0.09 | -0.012 (-0.039 to 0.014) | 0.55 |
| Bloating (0-100), mean (SD) | 23.0 (25.9) | 16.7 (21.8) | 15.8 (21.1) | 20.6 (27.0) | 19.4 (24.8) | 19.1 (22.1) | 13.5 (20.4) | 14.7 (21.1) | 15.9 (20.5) | 4.9 (0.2 to 9.6) | 0.039 | 9.3 (4.6 to 14.0) | <0.001 |
| Abdominal cramping (0-100), mean (SD) | 14.5 (21.0) | 12.4 (19.2) | 11.9 (18.7) | 12.3 (22.4) | 11.4 (17.9) | 13.5 (19.4) | 13.3 (22.4) | 13.4 (20.9) | 13.9 (21.1) | 3.5 (-1.1 to 8.0) | 0.13 | 3.0 (-1.6 to 7.6) | 0.20 |
| Flatulence (0-100), mean (SD) | 24.3 (25.1) | 20.6 (23.9) | 18.6 (22.0) | 24.5 (27.8) | 26.9 (27.0) | 28.7 (26.0) | 25.9 (24.5) | 21.6 (22.7) | 21.0 (20.2) | 9.0 (4.0 to 14.0) | <0.001 | 0.8 (-4.2 to 5.8) | 0.751 |
| Stool frequency, mean (SD) | 1.35 (0.70) | 1.45 (0.89) | 1.49 (0.92) | 1.30 (0.89) | 1.43 (0.76) | 1.54 (0.83) | 1.29 (0.82) | 1.43 (0.82) | 1.57 (0.87) | 0.1 (-0.1 to 0.3) | 0.370 | 0.2 (-0.1 to 0.4) | 0.142 |
| Stool consistency (BSC), mode (SD) | 3.64 (1.35) | 3.88 (1.27) | 3.77 (1.22) | 3.96 (1.11) | 3.90 (1.34) | 3.91 (1.21) | 3.76 (1.07) | 3.72 (1.13) | 3.79 (1.12) | -0.2 (-0.5 to 0.2) | 0.346 | -0.1 (-0.4 to 0.1) | 0.470 |
| Transit time in hours, mean (SD) | 27.0 (15.3) | 33.0 (21.0) | 34.9 (20.1) | 33.2 (22.6) | 30.3 (20.9) | 29.7 (18.7) | 31.2 (18.2) | 33.7 (18.7) | 31.1 (15.3) | -10.6 (-17.9 to -3.2) | 0.005 | -6.9 (-14.5 to 0.6) | 0.074 |

The treatment effects are expressed as slopes of the interaction effect (treatment\*time), tested using mixed model analysis. +Assessed by the MicrobeLink stool smear image analysis. \* $p < 0.05$ , \*\* $p < 0.01$ , and \*\*\* $p < 0.001$ for within-group changes from baseline assessed by Wilcoxon rank test on paired samples (for alpha diversity). Bloating: higher scores indicate more bloating. Abdominal cramping: higher scores indicate more cramping. Flatulence: higher scores indicate more flatulence. Abbreviations: ASVs, amplicon sequence variants; CG, control group; CI, confidence interval; HDF, high-dietary fiber; HFF, high-fermented food; PP, per protocol.

A similar effect was observed, as estimated by the MicrobeLink stool smear image analysis, showing a non-significant trend towards a lower alpha-diversity as well as an apparent increased Prevotella-to-Bacteroides (P/B) ratio in the HDF group relative to CG (Table 2). Beta diversity analysis (Bray–Curtis) indicated that between-subject dissimilarity decreased in all groups, with the greatest microbial community convergence observed in the HDF arm. The mean Bray–Curtis distance to the group centroid from baseline to week 8 decreased by 0.04 in the CG, 0.05 in HFF, and 0.08 in HDF (Figure S8), suggesting a more pronounced shift toward a uniform community structure in response to the high-fiber intervention.

Taxonomic compositional changes also differed by intervention. A multivariate classifier model based on the change in microbiota composition (week 8 minus week 0) identified distinct genera characterizing the HDF group versus the CG. In particular, increases in *Anaerostipes*, *Faecalibacterium*, and *Bifidobacterium* were most discriminative of the HDF intervention. BLAST-based 16S rRNA sequence annotation further pinpointed the species driving these genus-level changes: *Anaerostipes hadrus*, *Faecalibacterium prausnitzii*, and *Bifidobacterium adolescentis*, which are all species associated with a butyrogenic trophic network. These species showed the largest relative abundance gains in HDF compared to CG (Figure 5A,B). The classifier model distinguishing HDF from CG achieved an area under the ROC curve (AUC) of 0.81 ± 0.07, indicating strong predictive performance. Permutation testing confirmed that this performance was significantly better than chance (null model mean AUC ∼0.50 ± 0.13; permutation *p* < 0.001; Figure 5C). Independent, non-model-based, genus-level comparisons (Mann–Whitney U tests with false-discovery correction) corroborated the HDF-induced increases in *Anaerostipes*, *Faecalibacterium*, and *Bifidobacterium* (Q < 0.05 for each; Supplemental Excel File S2).

**Figure 5.**
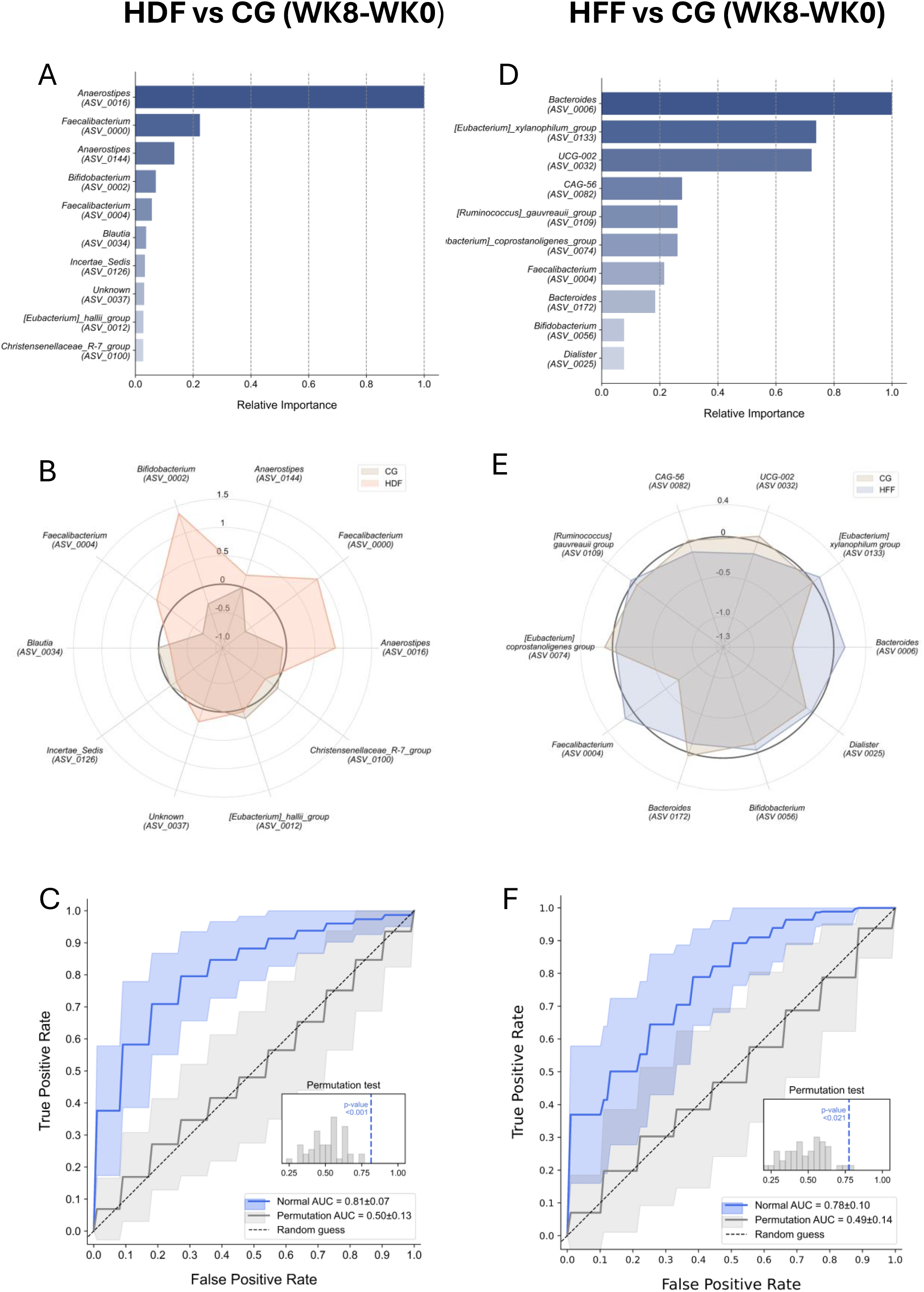
Top 10 predictive bacterial taxa and changes in their abundance comparing the end of intervention (week 8) to baseline (week 0) for high dietary fiber (HDF) and high fermented food (HFF) groups compared to the control group (CG). **A)** Bar plots showing relative importance of top ten predictive bacterial taxa for HDF *vs* HG. **B)** Radar plots of their changes in abundance for HDF *vs* CG. Species matching amplicon sequence variants: ASV_0016, *Anaerostipes hadrus*; ASV_0000, *Faecalibacterium prausnitzii*; ASV_0144, *Anaerostipes hadrus*; ASV_002, *Bifidobacterium adolescentis*; ASV_004, *Faecalibacterium duncaniae*; ASV_0034, *Blautia faecis*; ASV_0126, Ruminococcaceae sp.; ASV_0037, *Roseburia faecis*; ASV_0012, *Anaerobutyricum hallii* (p.k.a. *Eubacterium halii*); ASV_100, Christensenellacea sp. **C)** Model performance evaluation for HDF vs CG (WK8-WK0) using receiver operating characteristic (ROC) analysis, yielding an area under the curve (AUC). The inset shows the results of the permutation test was performed, generating a null distribution of AUC values. The observed model performance was indicated by the permutation *p*-value < 0.001) **D)** Bar plots showing relative importance of top ten predictive taxa HFF *vs* CG. **E)** Radar plots of their changes in abundance for HDF vs CG. Species matching amplicon sequence variants: ASV_0006, *Phocaeicola dorei*; ASV_0133, *Eubacterium xylanophilum*; ASV_0032, *Oscillospiraceae* sp; ASV_0082, *Hominisplanchenecus faecis*; ASV_109, *Lachnospiraceae* sp.; ASV_0074, Eubacteriaceae sp.; ASV_004, *Faecalibacterium duncaniae*; ASV_0172, *Phocaeicola massiliensis*. **F)** Model performance as in C) for HFF *vs* CG (WK8-WK0).

Compositional analysis using the same multi classifier model, also revealed distinct taxonomic shifts in the HFF group compared to the CG. Genera contributing to the observed separation included *Bacteroides*, *Ruminococcus*, and *Eubacterium* (Figure 5D–E). The BLAST-annotated species corresponding to the ASVs were *Phocaeicola dorei* (formerly *Bacteroides dorei*), *Eubacterium xylanophilum*, and *Ruminococcus gauvreauii*, taxa associated with saccharolytic fermentation and microbial cross-feeding. ROC analysis for the HFF group yielded an AUC of 0.78 ± 0.10, indicating moderate discriminatory performance. A permutation test confirmed that this classification was significantly better than expected by chance (mean permutation AUC = 0.49 ± 0.14; permutation *p*-value < 0.021; Figure 5F). However, in contrast to the HDF group, changes in the taxa were not statistically significant based on the non-model-based independent Mann–Whitney U tests, after correction for multiple comparisons (Supplemental excel file S2).

### 3.5 Shifts towards microbiota profiles associated with a younger age

Exploratory analysis using the multi classifier model identified ten microbial markers associated with age at baseline, across all treatment groups (*p* < 0.001), including higher abundances of amplicon sequence variants of *Blautia wexlerae* and *Agathobacter rectalis* (formerly known as *Eubacterium rectale*) in younger participants (≤50 years) (Figure 6A). Additionally, within the HFF group, changes in Shannon entropy over time differed by age, with participants over 50 years exhibiting a significantly greater increase in microbial diversity compared to individuals younger than 50 (β = 0.028, p < 0.05; Figure S9). To further assess the impact of the interventions on these age-related microbial species, we conducted a principal component analysis (PCA) exclusively using these ten marker species. The PCA visualization shows the microbiota composition differences between individuals younger and older than 50 years, with the centroids representing the delta shift from week 0 to week 8 (Figure S10). A greater negative delta indicates a more pronounced shift towards a microbiota profile of participants younger than 50 years at baseline, while a delta more closely to zero suggests a minimal change over time. From the bootstraps with 1,000 repetitions, the mean delta difference (D₂ - D₁) was calculated for each group, with the results demonstrating the strongest effect in the HFF group (-0.44), followed by HDF (-0.21) and CG (-0.19), indicating that the HFF intervention led to the most pronounced shift towards a microbiota composition associated with a younger age in our study population at baseline (Figure 6B).

**Figure 6.**
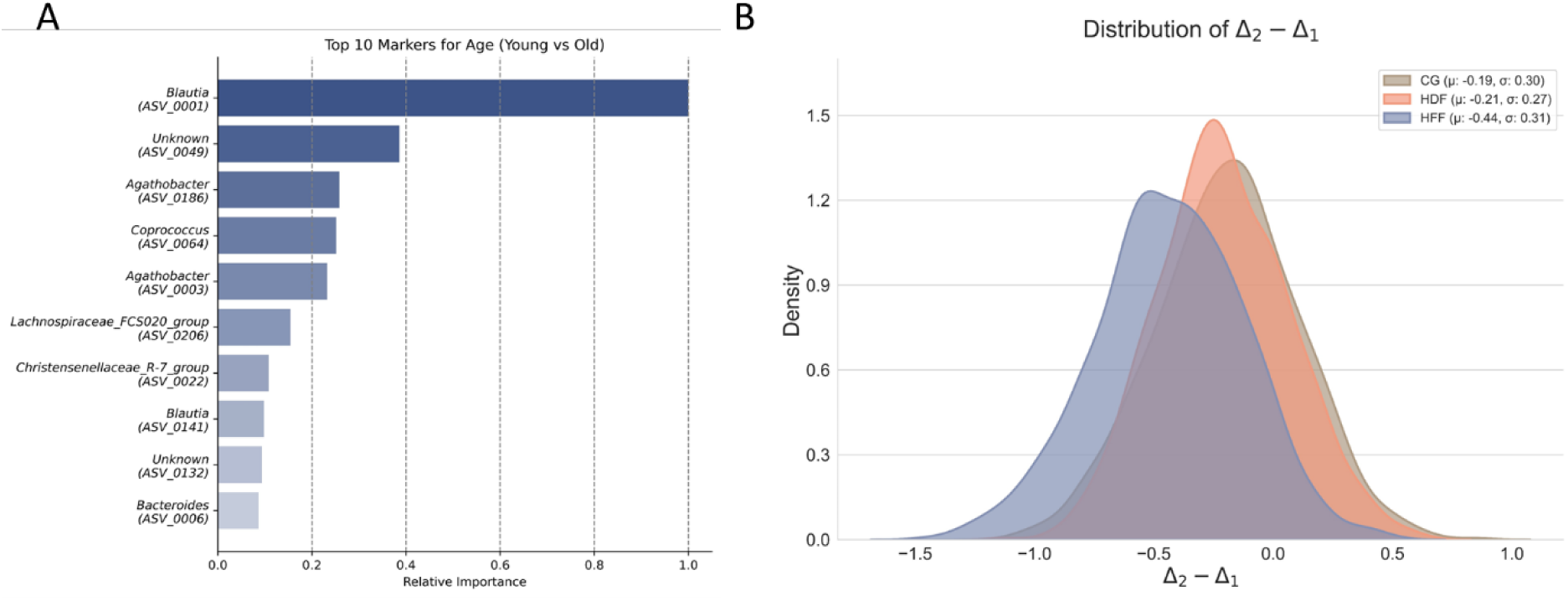
**A)** The top 10 microbial markers distinguishing younger (≤ 50 years old) from older adults (> 50 years old). **B)** Shifts in microbiota composition of participants > 50 years old towards a profile with taxa more prominent in study participants ≤ 50 years old (based on the top 10 markers). The mean delta difference (D₂ - D₁) was calculated for each group between week 8 and week 0. The corresponding PCA plots are displayed in Figure S10. BLAST species identification of amplicon sequence variants: ASV_0001, *Blautia wexlerae*; ASV_0049, *Ruminococcus bromii*; ASV_0186, *Agathobacter*; ASV_0064, *Coprococcus*; ASV_0003, *Agathobacter rectalis*.

### 3.6 Enhanced immunomodulatory effects in the HFF group

To assess changes in immune markers following the dietary interventions, effects on 92 inflammation-related protein markers were evaluated using delta values (week 8 minus week 0 and week 2 minus week 0). In the HFF group, five markers -CD8A, CD6, CD5 (related to T-cell activation), IL-18R1 (related to inflammatory signaling), and SIRT2 (related to longevity-associated deacetylase) were significantly upregulated compared to CG at week 8 (Q < 0.05; Figures 7, S11). No significant differences in immune markers were observed between the HDF and CG and at week 2 (Figure S12).

**Figure 7.**
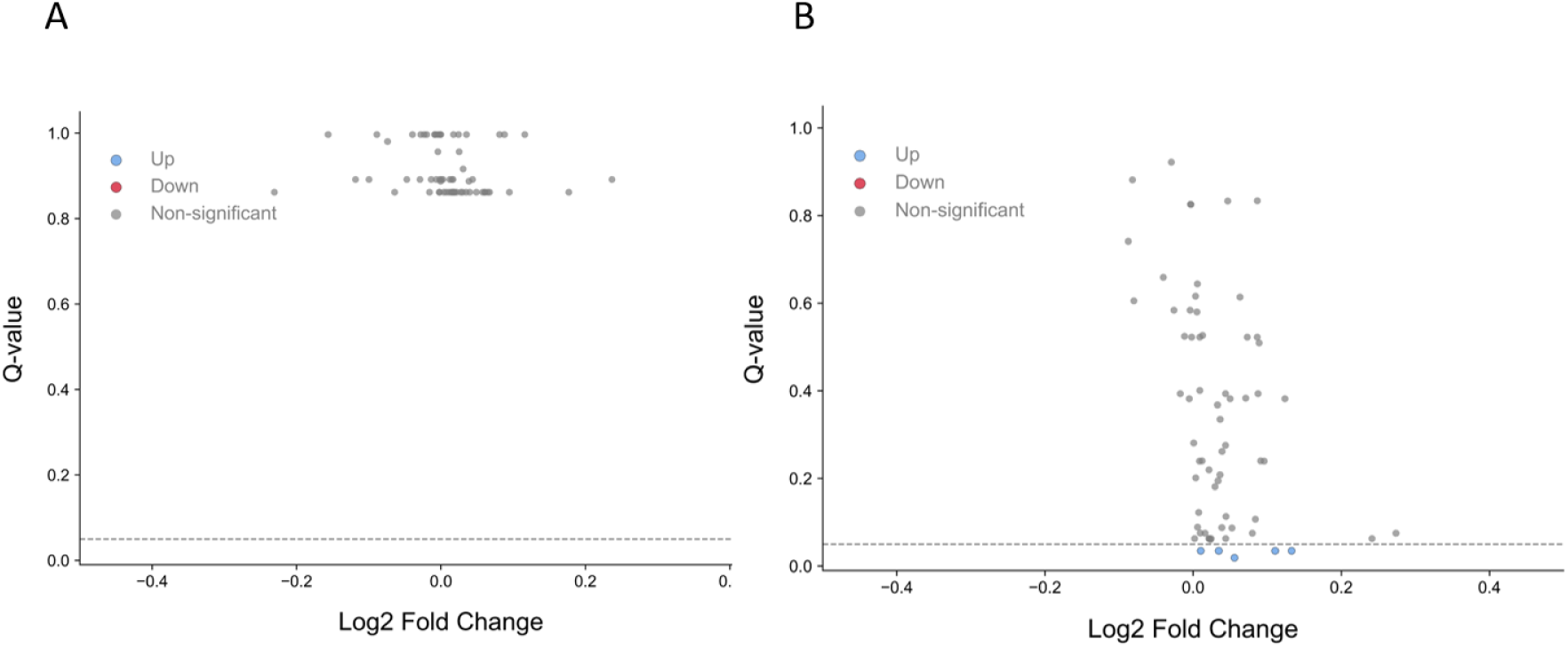
Volcano plots of Log2 fold change of blood inflammation markers of HDF versus CG (A) and HFF versus CG (B) between week 8 and week 0.

Within-group comparisons supported these findings: the HFF group showed substantial immune modulation between baseline and week 8, with 30 markers upregulated and 4 downregulated (Q < 0.05; Table S7). In contrast, the HDF group showed only two markers that significant changed over time: TNFRSF9 (related to T-cell co-stimulation) was upregulated, and TGF-α (related to epithelial proliferation) was downregulated. Both markers were also modulated in the HFF group. The CG exhibited no significant longitudinal changes in immune markers. Notably, no between-group differences were observed at week 2.

### 3.7 Transit time significantly decreased in the HDF group

A high inter-individual variation in transit time was observed across all groups, ranging from 3 to 126 hours. Changes in transit time were significantly different in the HDF group (Δ -222 min) compared to the CG (Δ +390 min). Indicating a significant reduction in transit time for the HDF group at both week 2 compared to week 0 (β = -480 min, *p*=0.03) and at week 8 compared to week 0 (β = -630 min, *p*=0.005) (Table 2), without affecting stool frequency and consistency. For the HFF group transit time (Δ -11 min), stool frequency and consistency remained stable over time. At baseline, exploratory analysis of microbial composition revealed distinct associations with gut transit time. Participants with a short transit time harbored higher abundances of saccharolytic taxa, including *Faecalibacterium* (butyrate-producing fiber fermenter) and *Prevotella* (plant polysaccharide degrader). In contrast, those with a long transit time were enriched in slow-growing, proteolytic or mucin-degrading taxa, such as *Methanobrevibacter* (methanogenesis), *Coprococcus* (branched-chain fatty acid production from amino acid fermentation), *Akkermansia* (mucus degradation), and members of the *Ruminococcaceae* family (protein and complex carbohydrate fermentation; Figure 8).

**Figure 8.**
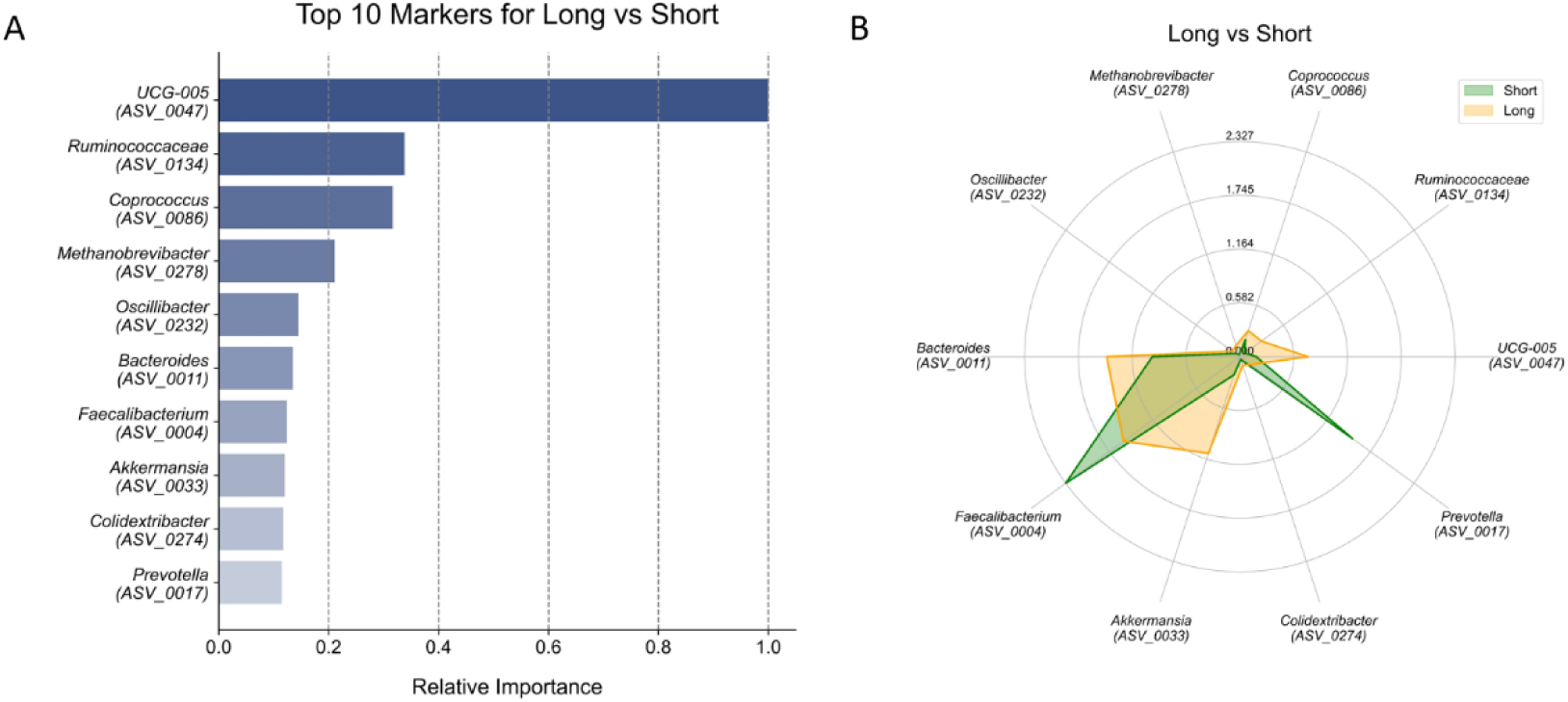
Top 10 genera changes in abundance comparing baseline (week 0) to end intervention (week 8) for long versus short transit time in all three study arms at baseline. Graphs show the 30% shortest versus the 30% longest transit time (≤23.7 and ≥33.8 hours). **A)** Bar plots show relative importance of top 8 bacterial taxa comparing end intervention (week 8) to baseline (week 0). **B)** Radar plots present how abundance of the top 10 bacterial genera or species differed (long versus short transit time). BLAST species identification of amplicon sequence variants: ASV_0047, Oscillospiraceae sp; ASV_0134, Ruminoccaceae sp.; ASV0086, *Coprococcus catus*; ASV_0278, *Methanobrevibacter smithii*; ASV_0232, Oscillospiraceae sp; ASV_0011, *Bacteroides uniformis*; ASV_004, *Faecalibacterium duncaniae*; ASV_0033, *Akkermansia muciniphila*; ASV_0274, *Colidextribacter*; ASV_0017, *Prevotella*.

### 3.8 Improvement in sleep quality in the HDF group

Sleep quality, as assessed using the Athens Insomnia Scale, significantly improved over time in the HDF group, compared to the change within the control group over the 8-week intervention period (β = -1.37, *p* = 0.03), with lower scores indicating better sleep (Table 3). Within the HDF group, sleep quality scores at follow-up were also significantly lower compared to baseline values (Δ = -0.9, *p* = 0.02), further supporting a sustained improvement in sleep quality after high fiber intake. Exploratory analysis revealed a trend toward a weak negative correlation between the change in fiber intake at follow-up and sleep scores within the group that followed the high dietary fiber diet during the intervention period (R = -0.3, *p* = 0.055), suggesting that increased fiber intake may be involved in improved sleep. Sleep quality did not change significantly in the HFF group compared to the control group.

**Table 3.** Secondary outcomes related to wellbeing, sleep and, perception and awareness of food choices at baseline (week 0), end of ramp-up (week 2), end of intervention (week 8) and follow-up (week 21) for the CG, HDF group and HFF group of the GEEF trial for the PP population.

|  | CG |  |  |  | HDF |  |  |  | HFF |  |  |  | Change between CG and HDF<br>delta week 8 and 0 |  | Change between CG and<br>HFF delta week 8 and 0 |  |
| --- | --- | --- | --- | --- | --- | --- | --- | --- | --- | --- | --- | --- | --- | --- | --- | --- |
| Week | 0 | 2 | 8 | 21 | 0 | 2 | 8 | 21 | 0 | 2 | 8 | 21 | Effect size (95% CI) | P-value | Effect size (95% CI) | P-value |
| DAQoL(0-9) | 0.9 (1.1) | 1.1 (1.0) | 0.9 (1.2) | 1.0 (1.0) | 0.8 (0.7) | 0.8 (0.8) | 0.9 (0.8) | 0.6 (0.7) | 0.7 (0.9) | 0.7 (1.0) | 0.9 (1.2) | 0.9 (1.3) | 0.04 (-0.38 to 0.47) | 0.84 | 0.13 (-0.34 to 0.52) | 0.55 |
| Sleep quality<br>(0-7) | 4.3 (3.7) | 4.5 (4.5) | 4.5 (3.8) | 3.9 (4.2) | 4.6 (3.0) | 4.1 (3.0) | 3.5 (3.0) | 3.7 (2.9) | 4.6 (3.9) | 4.7 (3.9) | 4.4 (3.7) | 4.2 (3.8) | -1.37 (-2.62 to -0.13) | **0.03** | -0.35 (-1.62 to 0.90) | 0.58 |
| Well-being<br>(0-100) | 64.8<br>(12.9) | 64.9<br>(15.0) | 65.2 (17.0) | 67.0 (15.2) | 64.5 (13.7) | 65.3 (13.1) | 66.5 (12.4) | 64.5 (11.7) | 66.5 (16.4) | 66.7 (16.9) | 66.4 (16.9) | 69.3 (14.8) | 1.67 (-3.66 to 6.99) | 0.54 | -0.52 (-5.91 to 4.87) | 0.85 |
| Subjective<br>knowledge<br>(5-35) | 20.3 (7.2) | 23.6 (6.2) | 23.5 (5.9) | 25.8 (5.2) | 19.6 (8.2) | 22.9 (7.1) | 24.0 (7.1) | 26.3 (5.6) | 21.1 (7.9) | 23.5 (6.2) | 24.0 (6.2) | 26.5 (5.5) | 1.24 (-0.66 to 3.14) | 0.21 | -0.31 (-2.78 to 1.07) | 0.75 |
| Dietary<br>intrinsic<br>motivation<br>(6-42) | 34.0 (6.6) | 34.7 (5.8) | 34.7 (6.0) | 33.4 (5.7) | 35.8 (5.1) | 35.2 (5.3) | 35.5 (5.0) | 35.4 (5.8) | 35.3 (5.3) | 34.6 (5.6) | 34.7 (5.7) | 34.8 (4.9) | -0.98 (-2.64 to 0.69) | 0.25 | -1.33 (-3.01 to 0.36) | 0.13 |
| Intention to<br>stay healthy<br>(3-21) | 17.9 (3.0) | 17.3 (2.5) | 17.6 (2.7) | 17.2 (2.8) | 18.3 (2.4) | 18.4 (2.7) | 18.3 (3.0) | 18.3 (2.5) | 18.6 (2.2) | 18.0 (2.5) | 18.0 (2.9) | 17.8 (2.1) | 0.23 (-0.78 to 1.24) | 0.66 | -0.32 (-1.34 to 0.70) | 0.54 |
| Self-regulation<br>(5-35) | 22.9 (6.1) | 23.3 (6.5) | 24.2 (5.8) | 23.2 (6.2) | 24.3 (4.9) | 25.4 (4.2) | 24.9 (4.8) | 24.5 (5.1) | 23.3 (5.4) | 23.7 (5.8) | 23.7 (5.8) | 24.0 (5.3) | -0.69 (-2.36 to 0.99) | 0.43 | -0.85 (-2.54 to 0.84) | 0.33 |
| Subjective<br>health (2-14) | 9.8 (2.1) | 9.9 (2.1) | 10.2 (2.0) | 9.9 (2.1) | 10.3 (1.7) | 11.1 (1.7) | 11.2 (1.6) | 10.6 (1.7) | 10.2 (2.0) | 10.7 (1.9) | 10.3 (2.1) | 10.1 (1.9) | 0.57 (-0.09 to 1.22) | 0.09 | -0.2 (-0.86 to 0.46) | 0.56 |
| Dietary self-efficacy (8-64) | 44.7 (7.0) | 44.3 (6.5) | 44.9 (6.5) | 43.8 (6.5) | 44.6 (6.9) | 46.8 (5.7) | 45.5 (6.5) | 46.0 (6.3) | 45.1 (7.4) | 44.5 (7.0) | 44.0 (6.4) | 44.1 (7.0) | 0.72 (-1.19 to 2.62) | 0.46 | -1.24 (-3.17 to 0.69) | 0.21 |

Data are presented as means (standard deviation). The treatment effect represents the interaction effect (treatment x time) based on mixed model analysis. \* $p < 0.05$ , \*\* $p < 0.01$ , and \*\*\* $p < 0.001$ for within-group changes from baseline assessed by paired t-test.
DQLQ: higher scores indicate worse digestion-associated quality of life. Sleep quality: lower scores indicate fewer sleep problems. Well-being: higher scores indicate better well-being. Subjective knowledge: higher scores indicate greater knowledge. Dietary intrinsic motivation: higher scores indicate greater motivation. Intention to stay healthy: higher scores indicate greater intention. Self-regulation: higher scores indicate better self-regulation. Subjective health: higher scores indicate better health. Dietary self-efficacy: higher scores indicate better self-efficacy. Abbreviations: CG, control group; DQLQ, digestion-associated quality of life questionnaire; HDF, high-dietary fiber; HFF, high-fermented food; PP, per protocol.

### 3.9 Trends in well-being and self-efficacy scores

For digestive-associated quality of life and well-being scores, no significant differences were observed between the treatment groups and the CG (Table 3). Across all three groups, knowledge scores improved throughout the ramp-up, intervention, and follow-up periods, though no significant differences were detected between the treatment groups and the CG. Scores for intrinsic motivation, intention to stay healthy, and total self-regulation remained stable over time and did not differ between the treatment groups and the CG (Table 3). Within the HDF group, subjective health and self-efficacy scores showed a slight but significant increase during the ramp-up period (β = 0.7, *p* = 0.04 and β = 2.7, *p* = 0.007, respectively) compared to changes in the CG. At follow-up, most participants indicated that they were still able to at least partly adhere to the dietary guidelines provided during the 8-week intervention. However, correlations between self-reported adherence at follow-up and actual changes in dietary fiber and fermented food intake were weak and not statistically significant (R = 0.12, *p* = 0.2 and R = 0.17, *p* = 0.06, respectively).

## 4. Discussion

This randomized dietary intervention study successfully increased the intake of dietary fiber and fermented foods, using a combination of dietary advice, study products and a citizen science (CS) approach. Our results demonstrate that integrating citizen science elements, such as education, self-monitoring, and sharing individual results, in a controlled trial is not only feasible but may have enhanced participant engagement and retention. These elements probably contributed to the sustained dietary changes observed at 21-week follow-up in both HDF and HFF intervention arms.

Although the implementation of the dietary recommendations was successful, no significant increase in alpha diversity was observed in either the HDF or HFF group compared to the CG. Interestingly, within-group increases in alpha diversity were found in the HFF and CG groups, but not in the HDF group. The absence of an alpha diversity increase in the HDF group is consistent with prior work indicating that an increase in fiber intake or supplementation does not consistently alter alpha diversity in healthy adults (Healey et al., 2017; So et al., 2018; Wastyk et al., 2021). This stability or decrease in alpha diversity may result from the ability of specific fibers to selectively promote certain functional groups while suppressing others, rather than increasing overall microbial diversity. If the interventions enrich specific beneficial functional groups and suppress non-beneficial groups, a lower alpha-diversity might still exert health-related benefits (Cantu-Jungles & Hamaker, 2023).

In line with this, distinct compositional shifts were evident in both intervention groups. The HDF intervention showed an increase in *Bifidobacterium* and *Anaerostipes* spp., which were shown to form a butyrogenic trophic chain from dried chicory root *in vitro* and to increase fecal and serum butyrate and other SCFA levels in two human intervention studies with the same study product (Omary et al., 2025; Puhlmann et al., 2022). *Bacteroides* species, which are abundantly present in the gut, degrade complex fibers into smaller oligosaccharides, which are then metabolized by *Bifidobacterium* into acetate and lactate, and subsequently utilized by *Anaerostipes* and *Faecalibacterium* for the production of butyrate, a SCFA linked to gut and systemic health, partly through their anti-inflammatory properties (Louis & Flint, 2017). Consistent with Puhlmann *et al*. (2022) and Omary et al (2025), the increase of butyrate-producing taxa observed in our study coincided with a decline in *Blautia*, associated with metabolic dysregulation (Liu et al., 2022).

In the HFF group, we observed microbiota changes that did not reflect direct colonization by microbes present in fermented foods, in agreement with previous findings (Wastyk et al., 2021). Rather, the intervention appeared to support the growth of resident saccharolytic taxa, including *Phocaeicola dorei*, *Eubacterium xylanophilum*, and *Ruminococcus gauvreauii*, likely through enhanced microbial cross-feeding. A novel finding was the age-dependent effect within the HFF group, participants over 50 years exhibited a more pronounced increase in Shannon diversity compared to younger participants. This is in line with earlier studies suggesting that the aging gut microbiota may be more responsive to dietary modulation, due to reduced ecological stability or altered host–microbiome interactions (Ghosh et al., 2020; Odamaki et al., 2016). Interestingly, at the end of the HFF intervention microbial profiles of older participants shifted toward a composition more similar to that of younger participants. This shift included an increased relative abundance of species such as *Blautia wexlerae* and *Agathobacter rectalis* (previously known as *Eubacterium rectale*), both of which have been associated with metabolic health and healthy ageing. Previously, administration of *Blautia wexlerae* ameliorated obesity and type 2 diabetes in mice via metabolic remodeling of the gut microbiota, notably stimulating butyrate-producers (Hosomi et al., 2022). In addition, *A. rectalis*, a butyrate producer, has been inversely associated with systemic inflammation and cognitive decline (Biagi et al., 2010; Lv et al., 2024).

The increase in alpha diversity within the CG was unexpected, given the absence of dietary changes. Participants received maltodextrin as a placebo and were only referred to general dietary guidelines. As maltodextrin intake was introduced in week 2, while the observed increase primarily occurred between week 0 and week 2 (Figure 4), it is unlikely that maltodextrin caused this increase in alpha diversity. Moreover, maltodextrin has been reported to exert limited and inconsistent effects on the gut microbiota (Almutairi et al., 2022), further supporting the involvement of an alternative underlying mechanism. Possibly, the observed increase in colonic transit time may have contributed to the diversity increase, which also occurred predominantly between week 0 and week 2 (Table 2). The slower gut transit time, specifically observed in the CG group, has been associated with greater microbial richness in other studies, likely due to longer substrate exposure and more stratified microenvironments (Vandeputte et al., 2016).

In contrast, participants in the HDF group reported faster transit times compared to CG. High-fiber diets are well-known to accelerate colonic transit, especially when administered in a high dose (>10 g/day) or for a long duration (>4 weeks) (de Vries et al., 2015). For example, wheat bran fiber has been shown to decrease gastrointestinal (GI) transit time while increasing stool weight (Muller-Lissner, 1988). This was consistent with our findings that the HDF group experienced a shorter bowel transit time, likely due to increased insoluble fiber intake (e.g., whole grains, legumes) acting as a bulking agent, or due to fermentable fibers increasing metabolite production, stimulating peristalsis. Shorter transit time can have positive downstream effects on the microbiota by reducing stagnation of stool and limiting over-fermentation in the colon, resulting in less proteolytic fermentation (Procházková et al., 2023). Additionally, improved GI motility often correlates with enhanced comfort and gut health, although this was not shown in our study.

A novel and striking finding in this study was the immunomodulatory effect observed in participants consuming high levels of fermented foods. By week 8, individuals in the HFF group showed significant increase in levels of five immune-related proteins in blood (Q < 0.05), including CD8A and CD6 (linked to cytotoxic and helper T-cell activation), IL-18R1 (involved in mucosal immunity and inflammatory signaling), CD5 (a negative regulator of T-cell activation), and SIRT2, a NAD⁺-dependent deacetylase associated with lymphocyte homeostasis, anti-inflammatory activity, and cellular longevity (De Oliveira et al., 2012; North et al., 2014; Ye et al., 2023). These effects differ from those reported by Wastyk et al. (2021), who observed reductions in 19 immune-related proteins following fermented food intake, using the same Olink platform methodology to measure 92 immune markers in blood. No significant changes were observed between groups during the first two weeks, indicating that the immunomodulatory effects at week 8 were probably not driven by fermented food intake alone. Instead, they most likely resulted from the fermentation-derived liquid supplement (AgeBiotic™), which was introduced at week 2 (Figure 7 and Figure S12). This supplement is rich in acetic acid and polyphenols, bioactive compounds with known immunomodulatory properties. Acetic acid, a short-chain fatty acid, is rapidly absorbed into the bloodstream and may influence immune markers detectable in blood (Qiu et al., 2019). Although Qiu et al. (2019) did not directly assess the markers identified in our study, their findings demonstrate that acetate enhances CD8⁺ T cell effector function, promotes histone acetylation, and boosts IFN-γ production under metabolic stress. These effects are consistent with the elevated levels of CD8A and CD6, while compensatory increases in CD5 and SIRT2 may reflect regulatory feedback and metabolic adaptation to enhanced T-cell activation. In addition, polyphenols can exert complementary effects by modulating oxidative stress and immune-related signaling cascades, as well as by shaping gut–immune interactions (Rodríguez-Daza et al., 2021; Shakoor et al., 2021). Together, these findings point to the AgeBiotic™ supplement as the most likely contributor to the immune marker changes observed in blood. However, effects of prolonged intake of fermented foods cannot be excluded, and a methodological difference — the use of dried blood spots, which capture both soluble proteins and cell-surface molecules from circulating immune cellscould have contributed to discrepancies with serum-based measurements reported in the Wastyk study.

Interestingly, the HDF intervention was associated with self-reported improved and sustained sleep quality, consistent with emerging evidence linking dietary fiber, the gut microbiome, and sleep via the gut–brain axis (Sen et al., 2021). Higher fiber intake has been correlated with increased time in slow-wave sleep and reduced nocturnal awakenings (St-Onge et al., 2016). Mechanistically, microbial fermentation of fiber produces SCFAs such as butyrate, which can modulate the synthesis of sleep-related neurotransmitters and gut hormones (e.g., serotonin, GLP-1, PYY), and attenuate inflammation, thereby influencing sleep architecture (Fusco et al., 2023). Increased levels of fecal and serum butyrate were found after dried chicory root intake in a trial with prediabetic subjects (Puhlmann et al., 2022). The observed sleep improvements in the HDF group may thus be mediated by enhanced SCFA production and gut–brain signaling. Additionally, high-fiber meals help stabilize blood glucose levels, preventing nocturnal glycemic fluctuations that can disrupt sleep (Goff et al., 2018). While further research is needed, these findings support a growing body of literature suggesting that a fiber-rich diet can positively influence sleep quality through microbiome-mediated pathways.

Another important finding is the modest, but significant improvement in dietary fiber and fermented food intake, three months post-intervention. Given the challenges with long-term dietary changes (Kumanyika et al., 2000), even modest improvements are a substantial achievement. This sustained change might be related to the incorporation of citizen science elements in the study (van de Put et al., 2024). Previous studies suggest that education, flexible recommendations and the allowance for self-monitoring via the provision of personal health data enhances behavior change (Michie et al., 2009; Middleton et al., 2013). Although the majority of participants reported feeling capable of adhering to the dietary advice at follow-up, this did not fully correlate with their actual dietary fiber and fermented food intake. This phenomenon is known as the intention-behavior gap (Conner & Norman, 2022). Factors moderating the relationship between intention and behavior include goal difficulty, moral norms, habits and temporal stability of intentions. Future dietary interventions could integrate action plans and uncertain rewards to further promote habit formation (Wood & Neal, 2016). Additionally, more personalized tools tailored to individual needs and preferences might help bridge the intention-behavior gap by providing targeted support and timely feedback.

This study has several strengths, including its randomized placebo-controlled design, relatively large sample size for a diet-microbiome trial, and comprehensive profiling of microbiome, immune, and health outcomes. The 21-week duration (including follow-up) is longer than many previous studies, providing insight into both immediate and medium-term effects of high-fiber and high-fermented food diets. The use of a citizen science framework is an innovative strength, as it likely contributed to the excellent compliance and sustained dietary behavior changes. The inclusion of dietary recommendations, in addition to the dietary products, allowed us to capture data in a more real-life setting. This approach is more aligned with how dietary products and supplements are typically recommended, as part of a dietary lifestyle intervention. The small, but sustained, dietary changes further enhance the translational relevance of the findings, it suggests that such diets are realistically implementable outside of a controlled research setting. Finally, by engaging participants directly through online platforms, social media, and local outreach, we were able to recruit individuals from across the Netherlands. This geographic diversity strengthens the generalizability of our results. Additionally, the multi-faceted outcome assessment, including microbiome sequencing, immune markers in blood, assessment of transit times, stool images, questionnaires about gut health, sleep, well-being, perception and awareness, and incorporation of machine learning methodology facilitates the identification of nonlinear associations and an integrated analysis of the systemic effects the dietary interventions in this study.

Despite these strengths, there are important limitations to acknowledge. First, although the sample size was decent, *i.e.* each of the three intervention arms had almost 50 participants, it may still have been underpowered to detect smaller changes in the microbiome or clinical markers. This might be partly due to the relative high number of excluded samples, caused by low read counts and missing baseline samples due to samples being lost in transit. Since analyses were performed on paired samples, participants with a missing sample at either time point had to be excluded entirely. Microbiome inter-individual variability is high, and as many other studies have noted, responses to a given diet differ based on one’s baseline microbiota (see *e.g.* Wang et al., 2025). A larger sample size could help stratify responders versus non-responders more robustly.

Second, the study population consisted of healthy, predominantly female, adults. Many of whom might have been health-conscious given the self-registration, although their intake of dietary fiber and fermented intake was relatively low. This limits generalizability to a broader and more diverse population: e.g. individuals with metabolic or inflammatory diseases, or those less motivated to change their diet, who might experience different effects. For example, we observed an increase in several immune-related markers following fermented food consumption. This may reflect an activation or modulation of immune function rather than a pro-inflammatory response, particularly in healthy individuals. In populations with higher baseline inflammation, such as those with metabolic or inflammatory diseases, the immunological effects of fermented foods might differ, potentially leading to reduction in inflammatory markers. Conversely, the fiber benefits might be dampened in people who lack key fiber-degrading bacteria, requiring a longer ramp-up period to effectively stimulate the growth of these gut bacteria.

Third, energy intake was reduced in both the CG and HDF groups at follow-up. While increased dietary fiber intake is associated with enhanced satiety and weight loss (Akhlaghi, 2024), it is unlikely that this caused the decreased energy intake observed in the present study, as body weight remained stable over time across all treatment groups. Therefore, the decrease in energy intake might be attributed to underreporting by participants, potentially due to reduced attention to dietary accuracy in the food diary three months post-intervention. Dietary adherence, while high, was self-reported and not blinded. Participants knew which diet they were following, so placebo effects or biases in reporting cannot be fully ruled out. We did include a control group with a placebo product, but the nature of dietary interventions makes blinding difficult. However, data-analysis was conducted in a blinded manner and objective measurements, including stool microbiota, blood markers, and transit time, provide confidence that the biological changes were real and diet-driven.

Another limitation is the lack of direct measurement of SCFA levels, gut permeability, metagenomes or immune cell function in this study. We infer mechanistic links (like SCFA-production) based on compositional data and known literature, but having measured fecal or plasma SCFA would strengthen the conclusions. However, these analyses are difficult to integrate in an at home study, as SCFA are volatile and therefore hard to capture in do-it-yourself kits. Similarly, functional immune assays (cytokine release, T-cell phenotyping) were not performed, so we rely on relative amounts of immune markers in blood as a proxy. Future studies should incorporate these outcomes, to confirm that the observed increased levels of for example CD8A translate to higher CD8 T-cell counts or activity. Finally, while the intervention was relatively long, it is unclear if the microbiome changes would persist after a prolonged dietary intervention. One might speculate that a partial regression of microbiome metrics will occur at follow-up, as participants largely return to their habitual diet, which is a common observation in dietary intervention studies.

These study findings provide insights for dietary recommendations aimed at promoting beneficial microbiota species, immune health and sleep quality. The citizen science-based approach demonstrated high feasibility and adherence, suggesting its potential applicability in upscaling of public health strategies. Policy implications include endorsing fiber-rich and fermented-food-rich diets as preventative measures to improve gut microbiome profiles and reduce systemic inflammation (Bell et al., 2017; Chilton et al., 2015). Public health programs should integrate educational components to encourage increased dietary awareness and long-term adherence to microbiome-supporting diets. More specifically, dietary fiber intake should be supported by substituting refined grains with whole grains and by encouraging fruit and nuts consumption, as participants were able to sustain the increased intakes in these categories.

Future studies should explore the long-term sustainability of microbiota, immune system and sleep improvements following dietary interventions rich in fiber and fermented foods. Specifically, research should assess whether continuous intake or intermittent periods of fermented food or high-fiber consumption are necessary for maintaining benefits. Further mechanistic studies employing direct measurement of SCFA-production, in particular acetic acid and butyric acid, gut barrier permeability, objective measures of sleep and functional immune responses (e.g., T-cell assays) will be critical to confirming hypothesized mechanistic pathways. Stratified analyses based on baseline microbiota and host factors such as age, metabolic health, and inflammatory status will also help identify responders versus non-responders and tailor dietary interventions accordingly. Lastly, large-scale population studies could validate the generalizability of the exploratory microbiota, sleep-related, and potential immune-related ‘rejuvenation’ effects seen here, extending to clinical applications, including gut-related diseases, or age-related inflammation.

In conclusion, our study’s findings are consistent with, and extend previous research, by showing that targeted high fiber or high fermented food dietary interventions can modulate the gut microbiome and thereby health factors in specific ways. We observed an increase in immune markers after fermented food intake and a faster transit time and improved sleep quality after increased dietary fiber intake, demonstrating the feasibility and value of engaging people in healthier food choices.

## Abbreviations

ACE: abundance-based coverage estimators index;
ASVs: amplicon sequence variants;
AUC: area under the curve;
BMI: body mass index;
CAZymes: carbohydrate-active enzymes;
CG: control group;
CS: citizen science;
GEEF: Gut health Enhancement by Eating favorable Food;
GI: gastrointestinal;
HDF: high-dietary fiber;
HFF: high-fermented food;
IQR: interquartile range;
ITT: intention to treat;
LMM: linear mixed model;
PP: per protocol;
RCT: randomized controlled trial;
SCFA: short-chain fatty acids.

## Ethical approval

Ethical approval was obtained from the Medical Ethical Committee Brabant in March 2023. The trial was prospectively registered at clinicaltrials.gov (NCT05900609) and conducted according to the declaration of Helsinki. Digital informed consent was obtained from each participant prior to inclusion in the study.

## Author contributions

Conceptualization: MvdB, MvdP, JPvS, NdW, RK; Methodology: MvdB, MvdP, NdW, RK; Data analysis: MvdB, DL, DM, EL; Provision of study materials: FvE, IR, NP, BB, MR; Conducting of research: MvdB, MvdP; Writing - Original Draft: MvdB, MvdP, RK; Writing - Review & Editing: MvdB, MvdP, EY, IBvdV, WMdV, NdW, RK; Data presentation: MvdB, DL, DM, EV; Supervision: NdW, RK; Funding acquisition: RK.

## Data availability

All data associated with this study are publicly available via Yoda, the research data management platform developed by Utrecht University, The Netherlands. The dataset can be accessed through the following DOI: https://doi.org/10.48338/VU01-K0YFNS. The dataset includes raw 16S rRNA gene sequencing data, O-link immune marker measurements, and questionnaire outcomes used to assess the distinct modulatory effects of high-fiber and fermented-food diets on gut microbiota composition, immune function, intestinal transit time, sleep quality, and overall well-being.

## Acknowledgements

We kindly acknowledge Odette Paling, Els Oosterink, Lonneke Janssen Duijghuijsen, Renee van den Hurk, Sara Hulscher, Nikkie Kaarsgaren, Saskia Meijboom from WUR FBR, Jenny Borkent, and Vincent Visker from WholeFiber for their support in the execution of the trial. Nicole de Roos for her critical review throughout the whole process. Ben Witteman for the medical advice during the study execution and critical review throughout the process, Arend Jan Copini (MDL Fonds) for support in project acquisition, Mark Bouwens (MDL Fonds) for support project management, Fons Voragen (Keep Food Simple) for support with study design, Javier Moreno (Horaizon) for support with the data analysis, Wilbert Sybesma (Microbiome Solutions) for stimulating discussions and connecting to WholeFiber and AniBiome, Fred Kaper (WholeFiber) for financial support, and emeritus Prof Ger Rijkers for critical review and interpretation of the immune outcomes.

## References

Akhlaghi, M. (2024). The role of dietary fibers in regulating appetite, an overview of mechanisms and weight consequences. Critical Reviews in Food Science and Nutrition, 64(10), 3139–3150. 10.1080/10408398.2022.2130160

Almutairi, R., Basson, A. R., Wearsh, P., Cominelli, F., & Rodriguez-Palacios, A. (2022). Validity of food additive maltodextrin as placebo and effects on human gut physiology: Systematic review of placebo-controlled clinical trials. European Journal of Nutrition, 61(6), 2853–2871. 10.1007/s00394-022-02802-5

Altmann, A., Toloşi, L., Sander, O., & Lengauer, T. (2010). Permutation importance: A corrected feature importance measure. Bioinformatics, 26(10), 1340–1347. 10.1093/bioinformatics/btq134

Altschul, S. F., Gish, W., Miller, W., Myers, E. W., & Lipman, D. J. (1990). Basic local alignment search tool. Journal of Molecular Biology, 215(3), 403–410. 10.1016/S0022-2836(05)80360-2

Armet, A. M., Deehan, E. C., O’Sullivan, A. F., Mota, J. F., Field, C. J., Prado, C. M., Lucey, A. J., & Walter, J. (2022). Rethinking healthy eating in light of the gut microbiome. Cell Host & Microbe, 30(6), 764–785. 10.1016/j.chom.2022.04.016

BBTools - DOE Joint Genome Institute. (2023). BBTools.

Bell, V., Ferrão, J., & Fernandes, T. (2017). Nutritional Guidelines and Fermented Food Frameworks. Foods, 6(8), 65.

Biagi, E., Nylund, L., Candela, M., Ostan, R., Bucci, L., Pini, E., Nikkïla, J., Monti, D., Satokari, R., Franceschi, C., Brigidi, P., & De Vos, W. (2010). Through Ageing, and Beyond: Gut Microbiota and Inflammatory Status in Seniors and Centenarians. PLoS ONE, 5(5), e10667. 10.1371/journal.pone.0010667

Bolyen, E., Rideout, J. R., Dillon, M. R., Bokulich, N. A., Abnet, C. C., Al-Ghalith, G. A., Alexander, H., Alm, E. J., Arumugam, M., Asnicar, F., Bai, Y., Bisanz, J. E., Bittinger, K., Brejnrod, A., Brislawn, C. J., Brown, C. T., Callahan, B. J., Caraballo-Rodríguez, A. M., Chase, J., … Caporaso, J. G. (2019). Reproducible, interactive, scalable and extensible microbiome data science using QIIME 2. Nature Biotechnology, 37(8), 852–857. 10.1038/s41587-019-0209-9

Bosco, N., & Noti, M. (2021). The aging gut microbiome and its impact on host immunity. Genes & Immunity, 22(5), 289–303. 10.1038/s41435-021-00126-8

Bowen, D. J. ; T. (1995). Controversies in Changing Dietary Behavior. In Nutrition and Health (1st Edition, pp. 45–66). CRC Press.

Callahan, B. J., McMurdie, P. J., Rosen, M. J., Han, A. W., Johnson, A. J. A., & Holmes, S. P. (2016). DADA2: High-resolution sample inference from Illumina amplicon data. Nature Methods, 13(7), 581–583. 10.1038/nmeth.3869

Cantu-Jungles, T. M., & Hamaker, B. R. (2023). Tuning Expectations to Reality: Don’t Expect Increased Gut Microbiota Diversity with Dietary Fiber. The Journal of Nutrition, 153(11), 3156–3163. 10.1016/j.tjnut.2023.09.001

Carding, S., Verbeke, K., Vipond, D. T., Corfe, B. M., & Owen, L. J. (2015). Dysbiosis of the gut microbiota in disease. Microb Ecol Health Dis, 26, 26191. 10.3402/mehd.v26.26191

Chen, S., Zhou, Y., Chen, Y., & Gu, J. (2018). fastp: An ultra-fast all-in-one FASTQ preprocessor. Bioinformatics, 34(17), i884–i890. 10.1093/bioinformatics/bty560

Chilton, S. N., Burton, J. P., & Reid, G. (2015). Inclusion of fermented foods in food guides around the world. Nutrients, 7(1), 390–404. 10.3390/nu7010390

Claesson, M. J., Jeffery, I. B., Conde, S., Power, S. E., O’Connor, E. M., Cusack, S., Harris, H. M. B., Coakley, M., Lakshminarayanan, B., O’Sullivan, O., Fitzgerald, G. F., Deane, J., O’Connor, M., Harnedy, N., O’Connor, K., O’Mahony, D., Van Sinderen, D., Wallace, M., Brennan, L., … O’Toole, P. W. (2012). Gut microbiota composition correlates with diet and health in the elderly. Nature, 488(7410), 178–184. 10.1038/nature11319

Clemente, J. C., Manasson, J., & Scher, J. U. (2018). The role of the gut microbiome in systemic inflammatory disease. *BMJ*, j5145. 10.1136/bmj.j5145

Conner, M., & Norman, P. (2022). Understanding the intention-behavior gap: The role of intention strength. Frontiers in Psychology, 13. 10.3389/fpsyg.2022.923464

De Oliveira, R. M., Sarkander, J., Kazantsev, A. G., & Outeiro, T. F. (2012). SIRT2 as a Therapeutic Target for Age-Related Disorders. Frontiers in Pharmacology, 3. 10.3389/fphar.2012.00082

Den Broeder, L., Devilee, J., Van Oers, H., Schuit, A. J., & Wagemakers, A. (2016). Citizen Science for public health. Health Promotion International, 33(3), 505–514. 10.1093/heapro/daw086

Ewels, P., Magnusson, M., Lundin, S., & Käller, M. (2016). MultiQC: summarize analysis results for multiple tools and samples in a single report. Bioinformatics, 32(19), 3047–3048. 10.1093/bioinformatics/btw354

Fabian Pedregosa, Gaël Varoquaux, Alexandre Gramfort, Vincent Michel, Bertrand Thirion, Olivier Grisel, Mathieu Blondel, Peter Prettenhofer, Ron Weiss, Vincent Dubourg, Jake Vanderplas, Alexandre Passos, David Cournapeau, Matthieu Brucher, Matthieu Perrot, & Édouard Duchesnay. (2011). Scikit-learn: Machine Learning in Python. J. Mach. Learn. Res., 12(null), 2825–2830.

Fusco, W., Lorenzo, M. B., Cintoni, M., Porcari, S., Rinninella, E., Kaitsas, F., Lener, E., Mele, M. C., Gasbarrini, A., Collado, M. C., Cammarota, G., & Ianiro, G. (2023). Short-Chain Fatty-Acid-Producing Bacteria: Key Components of the Human Gut Microbiota. Nutrients, 15(9), 2211. 10.3390/nu15092211

Garcia, S., Ordoñez, S., López-Molina, V. M., Lacruz-Pleguezuelos, B., Carrillo de Santa Pau, E., & Marcos-Zambrano, L. J. (2023). Citizen science helps to raise awareness about gut microbiome health in people at risk of developing non-communicable diseases. Gut Microbes, 15(1), 2241207. 10.1080/19490976.2023.2241207

Geurts, P., Ernst, D., & Wehenkel, L. (2006). Extremely randomized trees. Machine Learning, 63(1), 3–42. 10.1007/s10994-006-6226-1

Ghosh, T. S., Rampelli, S., Jeffery, I. B., Santoro, A., Neto, M., Capri, M., Giampieri, E., Jennings, A., Candela, M., Turroni, S., Zoetendal, E. G., Hermes, G. D. A., Elodie, C., Meunier, N., Brugere, C. M., Pujos-Guillot, E., Berendsen, A. M., De Groot, L. C. P. G. M., Feskins, E. J. M., … O’Toole, P. W. (2020). Mediterranean diet intervention alters the gut microbiome in older people reducing frailty and improving health status: The NU-AGE 1-year dietary intervention across five European countries. Gut, 69(7), 1218–1228. 10.1136/gutjnl-2019-319654

Goff, H. D., Repin, N., Fabek, H., El Khoury, D., & Gidley, M. J. (2018). Dietary fibre for glycaemia control: Towards a mechanistic understanding. Bioactive Carbohydrates and Dietary Fibre, 14, 39–53. 10.1016/j.bcdf.2017.07.005

Healey, G. R., Murphy, R., Brough, L., Butts, C. A., & Coad, J. (2017). Interindividual variability in gut microbiota and host response to dietary interventions. Nutrition Reviews, 75(12), 1059–1080. 10.1093/nutrit/nux062

Hosomi, K., Saito, M., Park, J., Murakami, H., Shibata, N., Ando, M., Nagatake, T., Konishi, K., Ohno, H., Tanisawa, K., Mohsen, A., Chen, Y.-A., Kawashima, H., Natsume-Kitatani, Y., Oka, Y., Shimizu, H., Furuta, M., Tojima, Y., Sawane, K., … Kunisawa, J. (2022). Oral administration of Blautia wexlerae ameliorates obesity and type 2 diabetes via metabolic remodeling of the gut microbiota. Nature Communications, 13(1), 4477. 10.1038/s41467-022-32015-7

Joos, R., Boucher, K., Lavelle, A., Arumugam, M., Blaser, M. J., Claesson, M. J., Clarke, G., Cotter, P. D., De Sordi, L., Dominguez-Bello, M. G., Dutilh, B. E., Ehrlich, S. D., Ghosh, T. S., Hill, C., Junot, C., Lahti, L., Lawley, T. D., Licht, T. R., Maguin, E., … Human Microbiome Action, C. (2024). Examining the healthy human microbiome concept. Nature Reviews Microbiology. 10.1038/s41579-024-01107-0

Kim, C. H. (2023). Complex regulatory effects of gut microbial short-chain fatty acids on immune tolerance and autoimmunity. Cellular & Molecular Immunology, 20(4), 341–350. 10.1038/s41423-023-00987-1

Kim, M., Oh, H.-S., Park, S.-C., & Chun, J. (2014). Towards a taxonomic coherence between average nucleotide identity and 16S rRNA gene sequence similarity for species demarcation of prokaryotes. International Journal of Systematic and Evolutionary Microbiology, 64(Pt_2), 346–351. 10.1099/ijs.0.059774-0

Köster, J., & Rahmann, S. (2012). Snakemake—A scalable bioinformatics workflow engine. Bioinformatics, 28(19), 2520–2522. 10.1093/bioinformatics/bts480

Kumanyika, S. K., Van Horn, L., Bowen, D., Perri, M. G., Rolls, B. J., Czajkowski, S. M., & Schron, E. (2000). Maintenance of dietary behavior change. Health Psychol, 19(1s), 42–56. 10.1037/0278-6133.19.suppl1.42

Leeuwendaal, N. K., Stanton, C., O’Toole, P. W., & Beresford, T. P. (2022). Fermented Foods, Health and the Gut Microbiome. Nutrients, 14(7), 1527.

Liu, S., Cao, R., Liu, L., Lv, Y., Qi, X., Yuan, Z., Fan, X., Yu, C., & Guan, Q. (2022). Correlation Between Gut Microbiota and Testosterone in Male Patients With Type 2 Diabetes Mellitus. Frontiers in Endocrinology, 13, 836485. 10.3389/fendo.2022.836485

Louis, P., & Flint, H. J. (2017). Formation of propionate and butyrate by the human colonic microbiota. Environmental Microbiology, 19(1), 29–41. 10.1111/1462-2920.13589

Lucassen, D. A., Brouwer-Brolsma, E. M., van de Wiel, A. M., Siebelink, E., & Feskens, E. J. M. (2021). Iterative Development of an Innovative Smartphone-Based Dietary Assessment Tool: Traqq (Vol. 169). 1940–087X.

Lv, X., Zhan, L., Ye, T., Xie, H., Chen, Z., Lin, Y., Cai, X., Yang, W., Liao, X., Liu, J., & Sun, J. (2024). Gut commensal Agathobacter rectalis alleviates microglia-mediated neuroinflammation against pathogenesis of Alzheimer disease. iScience, 27(11), 111116. 10.1016/j.isci.2024.111116

Makki, K., Deehan, E. C., Walter, J., & Bäckhed, F. (2018). The Impact of Dietary Fiber on Gut Microbiota in Host Health and Disease. Cell Host & Microbe, 23(6), 705–715. 10.1016/j.chom.2018.05.012

Marco, M. L., Heeney, D., Binda, S., Cifelli, C. J., Cotter, P. D., Foligné, B., Gänzle, M., Kort, R., Pasin, G., Pihlanto, A., Smid, E. J., & Hutkins, R. (2017). Health benefits of fermented foods: Microbiota and beyond. Current Opinion in Biotechnology, 44, 94–102. 10.1016/j.copbio.2016.11.010

Marco, M. L., Hill, C., Hutkins, R., Slavin, J., Tancredi, D. J., Merenstein, D., & Sanders, M. E. (2020). Should There Be a Recommended Daily Intake of Microbes? The Journal of Nutrition, 150(12), 3061–3067. 10.1093/jn/nxaa323

Michie, S., Abraham, C., Whittington, C., McAteer, J., & Gupta, S. (2009). Effective techniques in healthy eating and physical activity interventions: A meta-regression. Health Psychology, 28(6), 690–701. 10.1037/a0016136

Middleton, K. R., Anton, S. D., & Perri, M. G. (2013). Long-Term Adherence to Health Behavior Change. American Journal of Lifestyle Medicine, 7(6), 395–404. 10.1177/1559827613488867

Mohsen, A., Chen, Y.-A., Allendes Osorio, R. S., Higuchi, C., & Mizuguchi, K. (2022). Snaq: A Dynamic Snakemake Pipeline for Microbiome Data Analysis With QIIME2. Frontiers in Bioinformatics, 2. 10.3389/fbinf.2022.893933

Mölder, F., Jablonski, K. P., Letcher, B., Hall, M. B., Tomkins-Tinch, C. H., Sochat, V., Forster, J., Lee, S., Twardziok, S. O., Kanitz, A., Wilm, A., Holtgrewe, M., Rahmann, S., Nahnsen, S., & Köster, J. (2021). Sustainable data analysis with Snakemake. F1000Res, 10, 33. 10.12688/f1000research.29032.2

North, B. J., Rosenberg, M. A., Jeganathan, K. B., Hafner, A. V., Michan, S., Dai, J., Baker, D. J., Cen, Y., Wu, L. E., Sauve, A. A., Van Deursen, J. M., Rosenzweig, A., & Sinclair, D. A. (2014). SIRT 2 induces the checkpoint kinase BubR1 to increase lifespan. The EMBO Journal, 33(13), 1438–1453. 10.15252/embj.201386907

Odamaki, T., Kato, K., Sugahara, H., Hashikura, N., Takahashi, S., Xiao, J., Abe, F., & Osawa, R. (2016). Age-related changes in gut microbiota composition from newborn to centenarian: A cross-sectional study. BMC Microbiology, 16(1), 90. 10.1186/s12866-016-0708-5

Ojala, M., & Garriga, G. C. (2009). Permutation Tests for Studying Classifier Performance. 2009 Ninth IEEE International Conference on Data Mining, 908–913. 10.1109/ICDM.2009.108

Omary, L., Canfora, E. E., Puhlmann, M.-L., Gavriilidou, A., Rijnaarts, I., Holst, J. J., Op den Kamp-Bruls, Y. M. H., de Vos, W. M., & Blaak, E. E. (2025). Intrinsic chicory root fibers modulate colonic microbial butyrate-producing pathways and improve insulin sensitivity in individuals with obesity. *Cell Reports*. Medicine, 6(7), 102237. 10.1016/j.xcrm.2025.102237

Procházková, N., Falony, G., Dragsted, L. O., Licht, T. R., Raes, J., & Roager, H. M. (2023). Advancing human gut microbiota research by considering gut transit time. Gut, 72(1), 180–191. 10.1136/gutjnl-2022-328166

Puhlmann, M.-L., & de Vos, W. M. (2020). Back to the Roots: Revisiting the Use of the Fiber-Rich Cichorium intybus L. Taproots. Advances in Nutrition, 11(4), 878–890. 10.1093/advances/nmaa025

Puhlmann, M.-L., & de Vos, W. M. (2022). Intrinsic dietary fibers and the gut microbiome: Rediscovering the benefits of the plant cell matrix for human health. Frontiers in Immunology, 13. 10.3389/fimmu.2022.954845

Puhlmann, M.-L., Jokela, R., van Dongen, K. C. W., Bui, T. P. N., Hangelbroek, R. W. J. van, Smidt, H., de Vos, W. M., & Feskens, E. J. M. (2022). Dried chicory root improves bowel function, benefits intestinal microbial trophic chains and increases faecal and circulating short chain fatty acids in subjects at risk for type 2 diabetes. Gut Microbiome, 3, e4. 10.1017/gmb.2022.4

Qiu, J., Villa, M., Sanin, D. E., Buck, M. D., O’Sullivan, D., Ching, R., Matsushita, M., Grzes, K. M., Winkler, F., Chang, C.-H., Curtis, J. D., Kyle, R. L., Van Teijlingen Bakker, N., Corrado, M., Haessler, F., Alfei, F., Edwards-Hicks, J., Maggi, L. B., Zehn, D., … Pearce, E. L. (2019). Acetate Promotes T Cell Effector Function during Glucose Restriction. Cell Reports, 27(7), 2063–2074.e5. 10.1016/j.celrep.2019.04.022

Rijnaarts, I., de Roos, N. M., Wang, T., Zoetendal, E. G., Top, J., Timmer, M., Bouwman, E. P., Hogenelst, K., Witteman, B., & de Wit, N. (2021). Increasing dietary fibre intake in healthy adults using personalised dietary advice compared with general advice: A single-blind randomised controlled trial. Public Health Nutrition, 24(5), 1117–1128. 10.1017/S1368980020002980

Rijnaarts, I., de Roos, N., Zoetendal, E. G., de Wit, N., & Witteman, B. J. M. (2021). Development and validation of the FiberScreen: A short questionnaire to screen fibre intake in adults. J Hum Nutr Diet, 34(6), 969–980. 10.1111/jhn.12941

Rinninella, E., Raoul, P., Cintoni, M., Franceschi, F., Miggiano, G. A. D., Gasbarrini, A., & Mele, M. C. (2019). What is the Healthy Gut Microbiota Composition? A Changing Ecosystem across Age, Environment, Diet, and Diseases. Microorganisms, 7(1), 14.

Rodríguez-Daza, M. C., Pulido-Mateos, E. C., Lupien-Meilleur, J., Guyonnet, D., Desjardins, Y., & Roy, D. (2021). Polyphenol-Mediated Gut Microbiota Modulation: Toward Prebiotics and Further. Frontiers in Nutrition, 8, 689456. 10.3389/fnut.2021.689456

Ross, F. C., Patangia, D., Grimaud, G., Lavelle, A., Dempsey, E. M., Ross, R. P., & Stanton, C. (2024). The interplay between diet and the gut microbiome: Implications for health and disease. Nature Reviews Microbiology, 22(11), 671–686. 10.1038/s41579-024-01068-4

Roy, C. C., Kien, C. L., Bouthillier, L., & Levy, E. (2006). Short-chain fatty acids: Ready for prime time? Nutr Clin Pract, 21(4), 351–366. 10.1177/0115426506021004351

Sen, P., Molinero-Perez, A., O’Riordan, K. J., McCafferty, C. P., O’Halloran, K. D., & Cryan, J. F. (2021). Microbiota and sleep: Awakening the gut feeling. Trends in Molecular Medicine, 27(10), 935–945. 10.1016/j.molmed.2021.07.004

Shah, A. M., Tarfeen, N., Mohamed, H., & Song, Y. (2023). Fermented Foods: Their Health-Promoting Components and Potential Effects on Gut Microbiota. Fermentation, 9(2). 10.3390/fermentation9020118

Shakoor, H., Feehan, J., Apostolopoulos, V., Platat, C., Al Dhaheri, A. S., Ali, H. I., Ismail, L. C., Bosevski, M., & Stojanovska, L. (2021). Immunomodulatory Effects of Dietary Polyphenols. Nutrients, 13(3), 728. 10.3390/nu13030728

Shanahan, F., Ghosh, T. S., & O’Toole, P. W. (2021). The Healthy Microbiome-What Is the Definition of a Healthy Gut Microbiome? Gastroenterology, 160(2), 483–494. 10.1053/j.gastro.2020.09.057

Shivakoti, R., Biggs, M. L., Djoussé, L., Durda, P. J., Kizer, J. R., Psaty, B., Reiner, A. P., Tracy, R. P., Siscovick, D., & Mukamal, K. J. (2022). Intake and Sources of Dietary Fiber, Inflammation, and Cardiovascular Disease in Older US Adults. JAMA Netw Open, 5(3), e225012. 10.1001/jamanetworkopen.2022.5012

So, D., Whelan, K., Rossi, M., Morrison, M., Holtmann, G., Kelly, J. T., Shanahan, E. R., Staudacher, H. M., & Campbell, K. L. (2018). Dietary fiber intervention on gut microbiota composition in healthy adults: A systematic review and meta-analysis. The American Journal of Clinical Nutrition, 107(6), 965–983. 10.1093/ajcn/nqy041

St-Onge, M.-P., Roberts, A., Shechter, A., & Choudhury, A. R. (2016). Fiber and Saturated Fat Are Associated with Sleep Arousals and Slow Wave Sleep. Journal of Clinical Sleep Medicine, 12(01), 19–24. 10.5664/jcsm.5384

Tan, J., McKenzie, C., Potamitis, M., Thorburn, A. N., Mackay, C. R., & Macia, L. (2014). Chapter Three—The Role of Short-Chain Fatty Acids in Health and Disease. In F. W. Alt (Ed.), Advances in Immunology (Vol. 121, pp. 91–119). Academic Press.

van de Put, M., van den Belt, M., de Wit, N., & Kort, R. (2024). Rationale and design of a randomized placebo-controlled nutritional trial embracing a citizen science approach. Nutrition Research, 131, 96–110. 10.1016/j.nutres.2024.07.008

Van Hul, M., Cani, P. D., Petitfils, C., De Vos, W. M., Tilg, H., & El-Omar, E. M. (2024). What defines a healthy gut microbiome? Gut, 73(11), 1893. 10.1136/gutjnl-2024-333378

Vandeputte, D., Falony, G., Vieira-Silva, S., Tito, R. Y., Joossens, M., & Raes, J. (2016). Stool consistency is strongly associated with gut microbiota richness and composition, enterotypes and bacterial growth rates. Gut, 65(1), 57–62. 10.1136/gutjnl-2015-309618

Wang, L. S., Mo, Y. Y., Huang, Y. W., Echeveste, C. E., Wang, H. T., Chen, J., Oshima, K., Yearsley, M., Simal-Gandaraf, J., Battino, M., Xiao, J., Chen, J., Sun, C., Yu, J., & Bai, W. (2020). Effects of Dietary Interventions on Gut Microbiota in Humans and the Possible Impacts of Foods on Patients’ Responses to Cancer Immunotherapy. eFood, 1(4), 279–287. 10.2991/efood.k.200824.002

Wang, T., Holscher, H. D., Maslov, S., Hu, F. B., Weiss, S. T., & Liu, Y.-Y. (2025). Predicting metabolite response to dietary intervention using deep learning. Nature Communications, 16(1), 815. 10.1038/s41467-025-56165-6

Wastyk, H. C., Fragiadakis, G. K., Perelman, D., Dahan, D., Merrill, B. D., Yu, F. B., Topf, M., Gonzalez, C. G., Van Treuren, W., Han, S., Robinson, J. L., Elias, J. E., Sonnenburg, E. D., Gardner, C. D., & Sonnenburg, J. L. (2021). Gut-microbiota-targeted diets modulate human immune status. Cell, 184(16), 4137–4153.e14. 10.1016/j.cell.2021.06.019

Wood, W., & Neal, D. T. (2016). Healthy through Habit: Interventions for Initiating & Maintaining Health Behavior Change. Behavioral Science & Policy, 2(1), 71–83. 10.1177/237946151600200109

Yao, Y., Cai, X., Fei, W., Ye, Y., Zhao, M., & Zheng, C. (2022). The role of short-chain fatty acids in immunity, inflammation and metabolism. Critical Reviews in Food Science and Nutrition, 62(1), 1–12. 10.1080/10408398.2020.1854675

Ye, Y., Yang, K., Liu, H., Yu, Y., Song, M., Huang, D., Lei, J., Zhang, Y., Liu, Z., Chu, Q., Fan, Y., Zhang, S., Jing, Y., Esteban, C. R., Wang, S., Belmonte, J. C. I., Qu, J., Zhang, W., & Liu, G.-H. (2023). SIRT2 counteracts primate cardiac aging via deacetylation of STAT3 that silences CDKN2B. Nature Aging, 3(10), 1269–1287. 10.1038/s43587-023-00486-y

Yüksel, E., Voragen, A. G. J., & Kort, R. (2024). The pectin metabolizing capacity of the human gut microbiota. Critical Reviews in Food Science and Nutrition, 1–23. 10.1080/10408398.2024.2400235

Zheng, D., Liwinski, T., & Elinav, E. (2020). Interaction between microbiota and immunity in health and disease. Cell Research, 30(6), 492–506. 10.1038/s41422-020-0332-7

