## Supplemental Table and Figures for "Distinct modulatory effects of high-fiber and fermented-food diets on gut microbiota, immune function, transit time, and sleep quality in a citizen science randomized controlled trial"

### Supplementary Tables and Figures

**Table S1. Physical-chemical quantification (RU-MET-178) of AgeBiotics, fermentation-derived liquid supplement.** Presents the concentrations of targeted small molecules in AgeBiotics expressed in mol $L^{-1}$. Mass concentrations (mg $kg^{-1}$, equivalent to mg $L^{-1}$ assuming solution density ≈1 kg $L^{-1}$) were divided by each analyte's molar mass to yield molar concentrations. Total polyphenols cannot be converted to mol $L^{-1}$ due to their heterogeneous composition. This unified format facilitates direct comparison of ion, amino-acid, and phenolic-acid levels in absolute (molar) units. Participants in the HFF group consumed 57 mL daily (3 sachets)

| Analyte | Conc. (mg $kg^{-1}$) | Conc. (mol $L^{-1}$) |
|---|---|---|
| Acetic acid (E 260) | 27.398 | 0.456 |
| Total polyphenols | 7 000 | n.d.[1] |
| Sodium ($Na^+$) | 55.175 | $2.40 \times 10^{-3}$ |
| Calcium ($Ca^{2+}$) | 93.7 | $2.34 \times 10^{-3}$ |
| Magnesium ($Mg^{2+}$) | 320.35 | $1.32 \times 10^{-2}$ |
| GABA | 28.91 | $2.80 \times 10^{-4}$ |
| Caffeic acid | 68.59 | $3.81 \times 10^{-4}$ |
| Chlorogenic acid | 138.02 | $3.90 \times 10^{-4}$ |
| Quinic acid | 122.88 | $6.39 \times 10^{-4}$ |
| p-Hydroxybenzoic acid | 5.72 | $4.14 \times 10^{-5}$ |
| p-Coumaric acid | 1.03 | $6.28 \times 10^{-6}$ |
| Ferulic acid | 28.79 | $1.48 \times 10^{-4}$ |
| Salicylic acid | 17.46 | $1.26 \times 10^{-4}$ |
| Syringic acid | 397.29 | $2.00 \times 10^{-3}$ |
| Epicatechin | 562.51 | $1.94 \times 10^{-3}$ |

**Table S2. Targeted metabolomic of AgeBiotics, fermentation-derived liquid supplement profiling by capillary electrophoresis-Fourier transform mass spectrometry (CE-FTMS).** Participants in the HFF group consumed 57 mL daily (3 sachets).

| Analyte | Class | m/z | RT (min) | Peak Area |
|---|---|---|---|---|
| Spermidine | Polyamine | 146.165 | 3.6 | $3.3 \times 10^{-2}$ |
| Nicotinic acid (Vitamin $B_3$) | Pyridinium (Vit $B_3$) | 124.0393 | 7.84 | $3.2 \times 10^{-2}$ |
| Nicotinamide (Vitamin $B_3$) | Vitamin $B_3$ | 123.0552 | 5.86 | $3.2 \times 10^{-2}$ |

| | | | | |
|---|---|---|---|---|
| Nicotinamide riboside (Vitamin $B_3$) | Vitamin | 256.0813 | 9.73 | $4.2 \times 10^{-2}$ |
| Betaine | Quaternary ammonium | 118.0861 | 8.75 | $3.3 \times 10^{-1}$ |
| Glutathione (GSSG, oxidized) | Tripeptide | 307.0826 | 9.44 | $2.2 \times 10^{-1}$ |
| α-Tocopherol (Vitamin E) | Vitamin | 429.3733 | 14.2 | $3.0 \times 10^{-1}$ |
| Riboflavin (Vitamin $B_2$) | Vitamin | 377.1457 | 3.76 | $1.5 \times 10^{-1}$ |
| Epigallocatechin gallate (EGCG) | Polyphenol | 457.077 | 3.79 | $2.2 \times 10^{0}$ |
| Procyanidin B1 | Polyphenol | 579.1499 | 3.39 | $8.2 \times 10^{0}$ |
| Procyanidin B2 | Polyphenol | 579.1499 | 3.5 | $2.4 \times 10^{0}$ |
| Quercetin | Polyphenol | 301.0352 | 6 | $3.2 \times 10^{-2}$ |
| Gluconic Acid | Acid | 195.0509 | 7.34 | $4.5 \times 10^{0}$ |
| Lactic Acid | Acid | 89.0244 | 9.44 | $3.6 \times 10^{-1}$ |
| Malic Acid | Acid | 133.0142 | 16.25 | $3.1 \times 10^{0}$ |
| Citric Acid | Acid | 191.0197 | 18.58 | $6.0 \times 10^{0}$ |

**Table S3. SPIRIT diagram of study timeline and assessments of the GEEF trial.**

| Week | | -1 | 0 | 2 | 8 | | 21 |
|---|---|---|---|---|---|---|---|
| ENROLLMENT | | | | | | | |
| Screening questionnaire | x | | | | | | |
| Baseline characteristics | x | | | | | | |
| INTERVENTION | | | | | | | |
| High-dietary fiber | | | x | x | x | | |
| High-fermented foods | | | x | x | x | | |
| Control | | | x | x | x | | |
| ASSESSMENTS | | | | | | | |
| Body weight | | x | | x | x | | x |
| Fecal sample | | x (2) | | x | x (2) | | |
| Stool smear image | | x (2) | | x | x (2) | | |
| Dried blood spots | | x (2) | | x | x (2) | | |
| Blue muffins | | x | | x | x | | |
| Food diary | | x | | x | x | | x |
| Sleep quality | | x | | x | x | | x |
| DQLQ | | x | | x | x | | x |
| Wellbeing | | x | | x | x | | x |
| Perception and awareness | | x | | x | x | | x |
| FFF | | x | | x | x | | |
| Stool frequency | | | | | | x | |
| Stool consistency | | | | | | x | |

| | | | | |
|---|---|---|---|---|
| GI complaints | | x | | |
| Compliance | | x | | |
| Adverse events | | | x | |
| Supplement use | | | x | |
| Medication use | | | x | |
| Evaluation dietary advice | x | | | |
| Evaluation | | | | x |

[a] Baseline measurement. [b] End ramp-up measurement. [c] Endline measurement. [d] Follow-up measurement.
Abbreviations: DQLQ, digestion-associated quality of life questionnaire; FFF, fermented foods frequency questionnaire; GI, gastrointestinal

**Table S4. Per protocol population of week eight-zero pairs for each outcome compared to total number of included participants.**

| Outcome | CG | Reasons | HDF | Reasons | HFF | Reasons |
|---|---|---|---|---|---|---|
| Gut microbiota | 35/48 | Non-compliance dietary intervention n=1<br>Missing week 0 or week 8 sample n=9<br>Technical error 16S rRNA V3-V4 analysis n=3 | 33/49 | <75% completion questionnaires n=1<br>Antibiotic use n=1<br>Drop-out n=1<br>Non-compliance dietary intervention n=3<br>Missing week 0 or week 8 sample n=10 | 31/49 | <75% completion questionnaires n=3<br>Antibiotic use n=1<br>Drop-out n=1<br>Non-compliance dietary intervention n=3<br>Missing week 0 or week 8 sample n=10 |
| Inflammatory protein biomarkers | 46/48 | Non-compliance dietary intervention n=1<br>Missing week 0 or week 8 sample n=1 | 40/49 | <75% completion questionnaires n=1<br>Antibiotic use n=1<br>Drop-out n=1<br>Non-compliance dietary intervention n=3<br>Missing 0 or week 8 sample n=3 | 41/49 | <75% completion questionnaires n=3<br>Antibiotic use n=1<br>Drop-out n=1<br>Non-compliance dietary intervention n=3 |
| Dietary intake | 47/48 | Non-compliance dietary intervention n=1 | 43/49 | <75% completion questionnaires n=1<br>Drop-out n=1<br>Incomplete data n=1<br>Non-compliance dietary intervention n=3 | 40/49 | <75% completion questionnaires n=3<br>Drop-out n=1<br>Incomplete data n=2<br>Non-compliance dietary intervention n=3 |
| Stool frequency and consistency | 44/48 | Missing data n=3<br>Non-compliance dietary intervention n=1 | 41/49 | <75% completion questionnaires n=1<br>Drop-out n=1<br>Missing data=3<br>Non-compliance dietary intervention n=3 | 41/49 | <75% completion questionnaires n=3<br>Drop-out n=1<br>Non-compliance dietary intervention n=3 |
| Questionnaire outcomes | 48/48 | Not applicable | 48/49 | Drop-out n=1 | 48/49 | Drop-out n=1 |
| Body weight | 47/48 | Non-compliance dietary intervention n=1 | 44/49 | <75% completion questionnaires n=1<br>Drop-out n=1<br>Non-compliance dietary intervention n=3 | 42/49 | <75% completion questionnaires n=3<br>Drop-out n=1<br>Non-compliance dietary intervention n=3 |

Abbreviations: CG, control group; HDF, high-dietary fiber; HFF, high-fermented food.

**Table S5. Overview of all adverse events reported for the HDF group, HFF group and CG during the 8-week study period of the GEEF trial.**

| | CG | HDF | HFF | Total |
|---|---|---|---|---|
| Flu/corona/fever, n (%) | 6 (26.1) | 5 (21.7) | 12 (52.2) | 23 (100.0) |
| Common cold, n (%) | 10 (23.3) | 17 (39.5) | 16 (37.2) | 43 (100.0) |
| Gastrointestinal complaints, n (%) | 14 (42.4) | 7 (21.2) | 12 (36.4) | 33 (100.0) |
| Menstrual complaints, n (%) | 27 (47.4) | 20 (35.1) | 10 (17.5) | 57 (100.0) |
| Headache/tiredness, n (%) | 27 (31.4) | 30 (34.9) | 29 (33.7) | 86 (100.0) |
| Complaints related to study sachets*, n (%) | 4 (33.3) | 1 (8.3) | 7 (58.3) | 12 (100.0) |
| Other complaints, n (%) | 12 (24.0) | 17 (34.0) | 21 (42.0) | 50 (100.0) |
| Supplement use, n (%) | 10 (38.5) | 5 (19.2) | 11 (42.3) | 26 (100.0) |
| Medication use, n (%) | 12 (26.7) | 8 (17.8) | 25 (55.6) | 45 (100.0) |
| Vaccination, n (%) | 5 (41.7) | 3 (25.0) | 4 (33.3) | 12 (100.0) |

*Participants specifically mentioned the study product as the presumed cause of the complaint, all concerned mild gastrointestinal complaints. Abbreviations: CG, control group; HDF, high-dietary fiber; HFF, high-fermented food.

**Table S6. Mean dietary intake per day excluding consumption of study products for the HDF group, HFF group and CG at baseline (week 0), end of ramp-up (week 2), end of intervention (week 8) and end of follow-up (week 21) for the PP population. Values show means with standard deviations in parentheses.**

| | **CG** | | | | **HDF** | | | | **HFF** | | | |
|---|---|---|---|---|---|---|---|---|---|---|---|---|
| **Week** | 0 | 2 | 8 | 21 | 0 | 2 | 8 | 21 | 0 | 2 | 8 | 21 |
| Number of participants | 47 | 46 | 47 | 46 | 43 | 43 | 43 | 42 | 40 | 40 | 40 | 40 |
| Energy (kcal) | 1995.7 (488.4) | 1943.4 (458.0) | 1920.9 (518.9) | 1841.2* (461.9) | 1878.6 (472.5) | 2008.0 (562.4) | 1894.9 (542.1) | 1659.0* (412.3) | 1927.6 (458.1) | 1901.5 (362.4) | 1991.5 (523.6) | 1805.2 (524.7) |
| Dietary fiber (g) | 22.1 (7.5) | 21.1 (6.3) | 20.4 (6.3) | 20.8 (6.0) | 20.4 (6.2) | 30.2† (8.0) | 30.1† (7.7) | 21.5 (6.5) | 21.2 (7.9) | 19.6 (7.8) | 21.2 (9.1) | 20.1 (8.9) |
| CH (g) | 211.6 (60.8) | 203.4 (61.8) | 199.8 (66.4) | 188.8 (55.5) | 195.5 (52.7) | 208.5 (54.7) | 196.5 (55.0) | 176.3 (57.1) | 196.6 (56.7) | 195.1 (45.5) | 202.3 (59.2) | 184.0 (65.8) |
| Protein (g) | 76.9 (22.2) | 74.9 (21.4) | 73.3 (20.0) | 72.3 (19.5) | 67.2 (20.9) | 73.5 (25.9) | 66.9 (20.7) | 62.3 (16.9) | 71.1 (19.5) | 76.6 (18.2) | 79.4† (24.6) | 73.8 (24.6) |
| Fat (g) | 82.8 (27.8) | 81.1 (25.8) | 79.4 (28.0) | 79.3 (27.1) | 78.6 (25.1) | 82.4 (27.4) | 79.7 (32.8) | 67.9 (21.1) | 80.9 (22.6) | 78.1 (20.0) | 83.4 (29.2) | 74.6 (23.4) |
| Saturated fat (g) | 28.0 (11.1) | 26.9 (9.5) | 26.1 (11.5) | 28.5 (13.2) | 24.8 (9.0) | 23.5 (9.4) | 22.5 (10.0) | 22.0 (7.3) | 26.3 (8.2) | 25.6 (7.6) | 30.0† (12.6) | 25.5 (9.6) |
| Natrium (mg) | 2111.5 (748.9) | 2020.7 (685.2) | 2058.6 (829.4) | 2063.3 (761.6) | 1942.0 (552.7) | 1804.5 (540.5) | 1777.4 (835.8) | 1796.0 (572.6) | 2219.4 (701.2) | 2390.9 (568.7) | 2487.1 (923.8) | 2199.8 (928.9) |
| FF (portions) | 0.7 (0.7) | 0.8 (0.8) | 0.8 (0.8) | 1.0 (1.1) | 0.6 (0.6) | 0.6 (0.6) | 0.6 (0.7) | 0.8 (1.0) | 0.8 (0.7) | 4.1† (1.2) | 4.3† (1.3) | 1.4* (1.0) |

† denotes significant difference of p=<0.05 compared to change in control group based on linear mixed model analysis.

* p < 0.05, ** p < 0.01, and *** p < 0.001 for within-group changes from baseline assessed by paired t-test.

Abbreviations: CG, control group; CH, carbohydrates; FF, fermented foods; g, grams; HDF, high-dietary fiber; HFF, high-fermented food; ITT, intention to treat; kcal, kilocalories; mg, milligrams

**Table S7 Upregulated (red) and downregulated (green) immune markers in dried blood spots.**

| between group analysis | | | | within group analysis | | | | | |
|---|---|---|---|---|---|---|---|---|---|
| HDF *versus* CG | | HFF *versus* CG | | CG | | HDF | | HFF | |
| Q<0.1 | Q<0.05 | Q<0.1 | Q<0.05 | Q<0.1 | Q<0.05 | Q<0.1 | Q.<0.05 | Q<0.1 | Q<0.05 |
| | | | | TGF α | | TGF α | TGF α | TGF α | TGF α |
| | | | | CASP 8 | | | | CASP 8 | CASP 8 |
| | | | | | | | | EN_RAGE | EN_RAGE |
| | | | | | | | | MCP 1 | MCP 1 |
| | | | | | | TNFRSF9 | TNFRSF9 | TNFRSF9 | TNFRSF9 |
| | | CD8A | CD8A | | | | | CD8A | CD8A |
| | | CD6 | CD6 | | | | | CD6 | CD6 |
| | | IL_18R1 | IL_18R1 | | | | | IL_18R1 | IL_18R1 |
| | | CD5 | CD5 | | | | | CD5 | CD5 |
| | | SIRT2 | SIRT2 | | | | | SIRT2 | SIRT2 |
| | | TNF | | | | | | TNF | TNF |
| | | CXCL6 | | | | | | CXCL6 | CXCL6 |
| | | IL8 | | | | | | IL8 | IL8 |
| | | VEGFA | | | | | | VEGFA | VEGFA |
| | | CD244 | | | | | | CD244 | CD244 |
| | | AXIN1 | | | | | | AXIN1 | AXIN1 |
| | | OSM | | | | | | OSM | OSM |
| | | CXCL1 | | | | | | CXCL1 | CXCL1 |
| | | CXCL5 | | | | | | CXCL5 | CXCL5 |
| | | PD_L1 | | | | | | PD_L1 | PD_L1 |
| | | HGF | | | | | | HGF | HGF |
| | | CCL28 | | | | | | CCL28 | CCL28 |
| | | CD40 | | | | | | CD40 | CD40 |
| | | STAMBP | | | | | | STAMBP | STAMBP |
| | | | | | | | | MMP_1 | MMP_1 |
| | | | | | | | | CCL19 | CCL19 |
| | | | | | | | | IL_15RA | IL_15RA |
| | | | | | | | | IL7 | IL7 |
| | | | | | | | | OPG | OPG |
| | | | | | | | | MCP_4 | MCP_4 |
| | | | | | | | | CCL11 | CCL11 |
| | | | | | | | | CCL25 | CCL25 |
| | | | | | | | | CSF_1 | CSF_1 |
| | | | | | | | | ADA | ADA |
| | | | | | | | | CDCP1 | |
| | | | | | | | | CST5 | |
| | | | | | | | | SCF | |
| | | | | | | | | MMP_10 | |
| | | | | | | | | CCL23 | |
| | | | | | | | | Flt3L | |

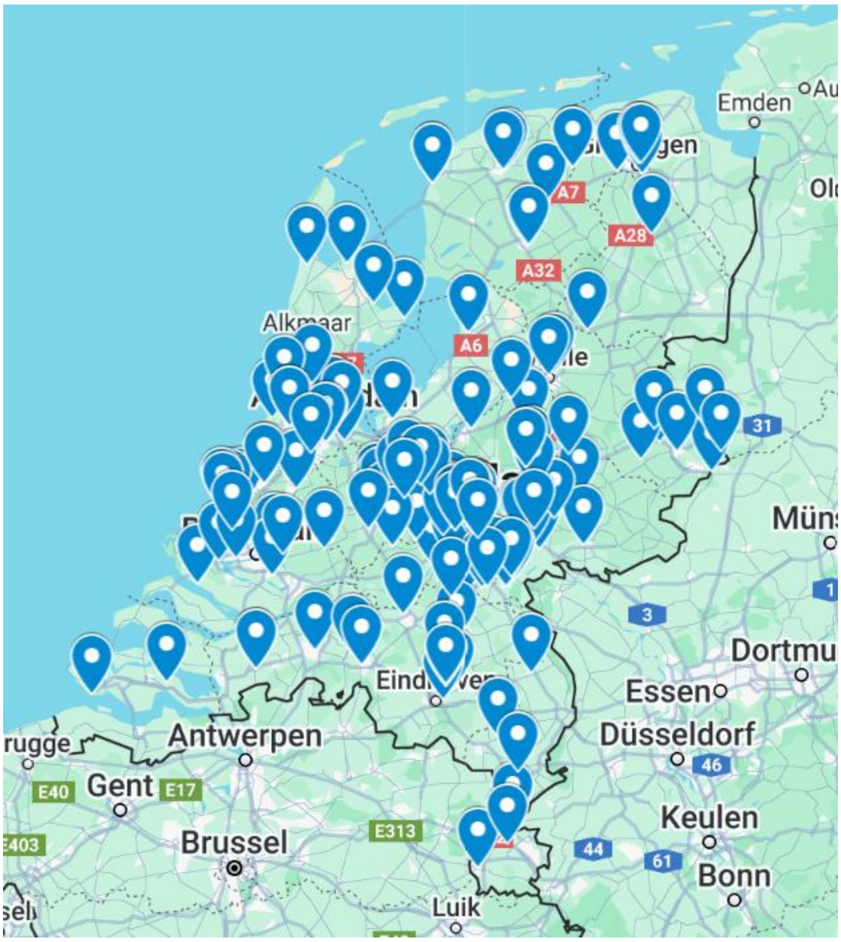


**Figure S1. Geographical spread of participants across the Netherlands**

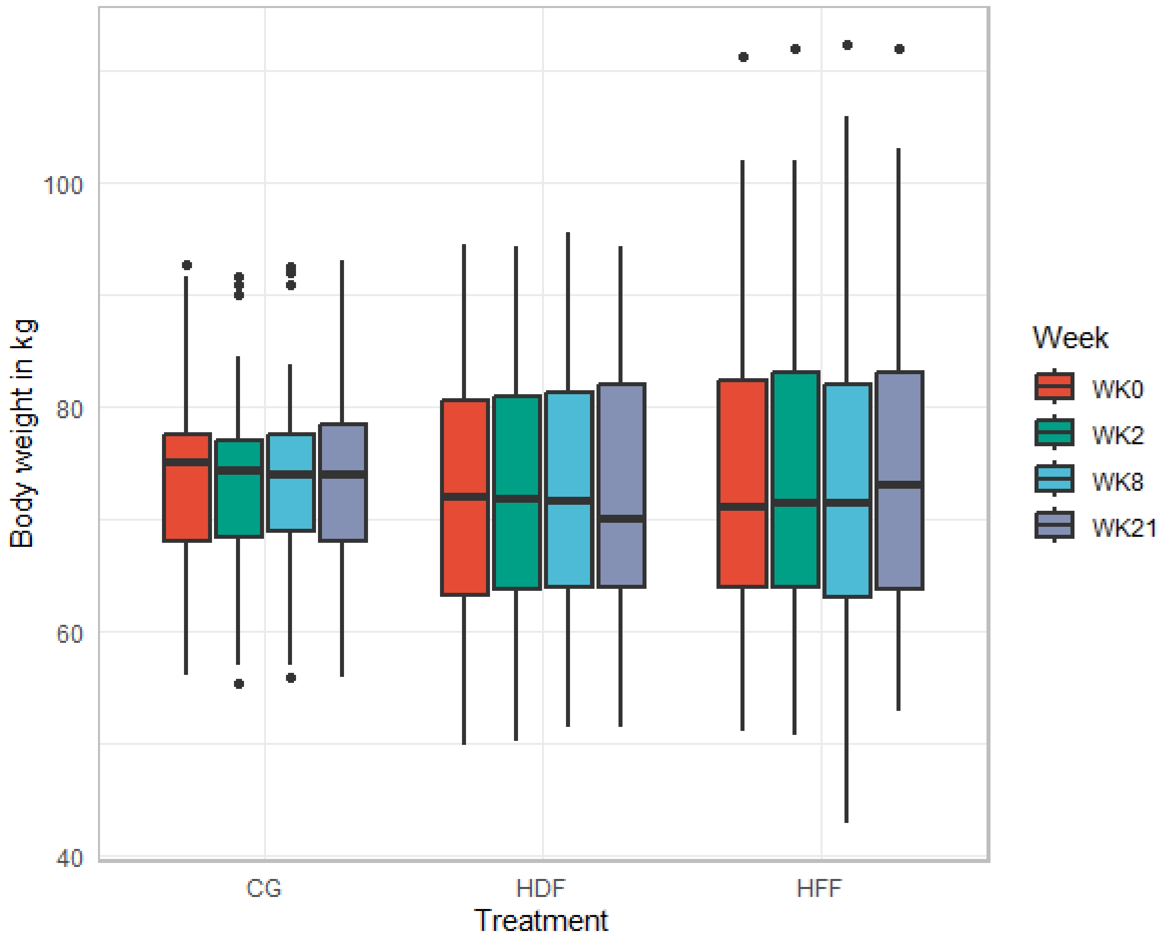


**Figure S2. Body weight in kilograms at baseline (week 0), end of ramp-up (week 2), end of intervention (week 8) and end of follow-up (week 21), for the PP population.** Abbreviations: CG, control group; HDF, high-dietary fiber; HFF, high-fermented food; PP, per protocol.

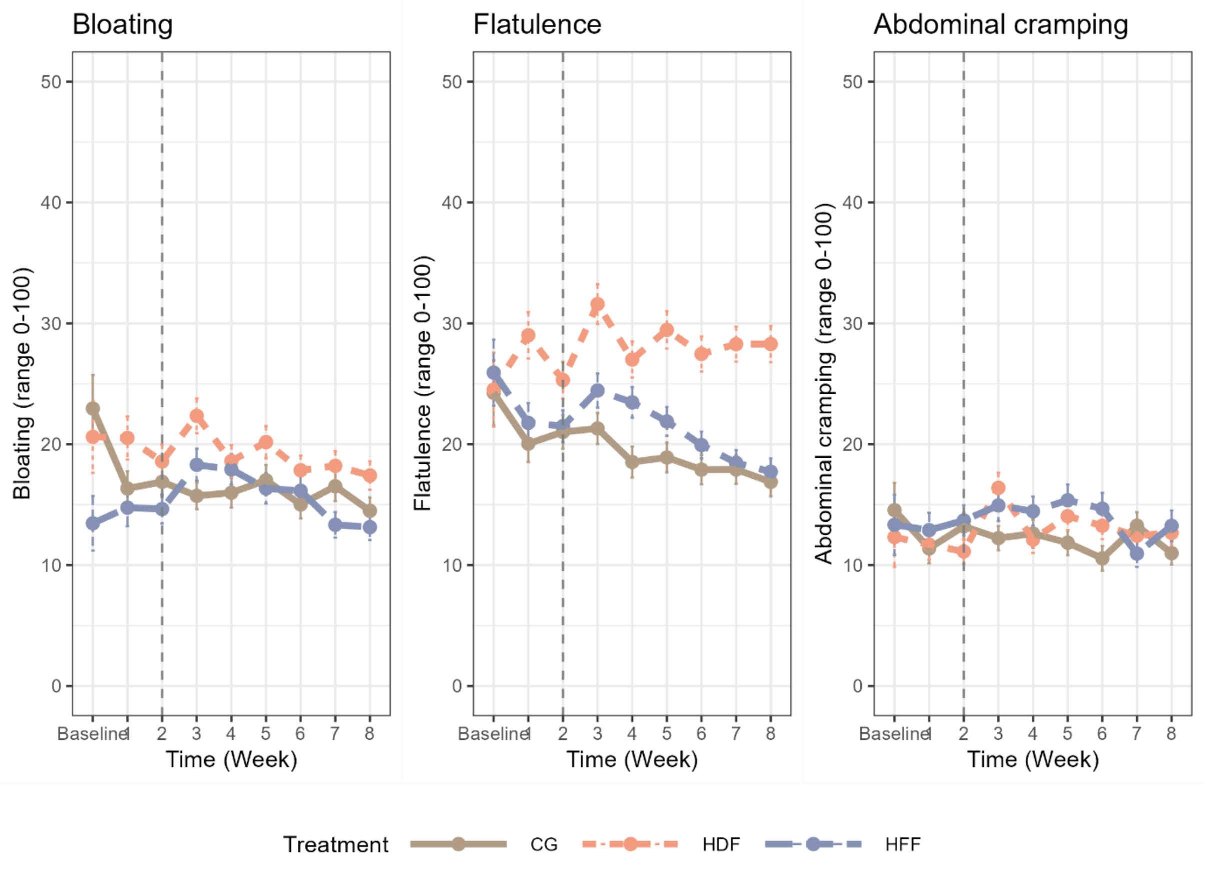


**Figure S3. Average bloating (figure A), flatulence (figure B), and abdominal cramping (figure C) complaints per week for the CG, HDF group and HFF group of the GEEF trial, for the PP population.** Baseline values show day 0-2, the vertical gray line represents the end of the ramp-up period. Abbreviations: CG, control group; HDF, high-dietary fiber; HFF, high-fermented food.

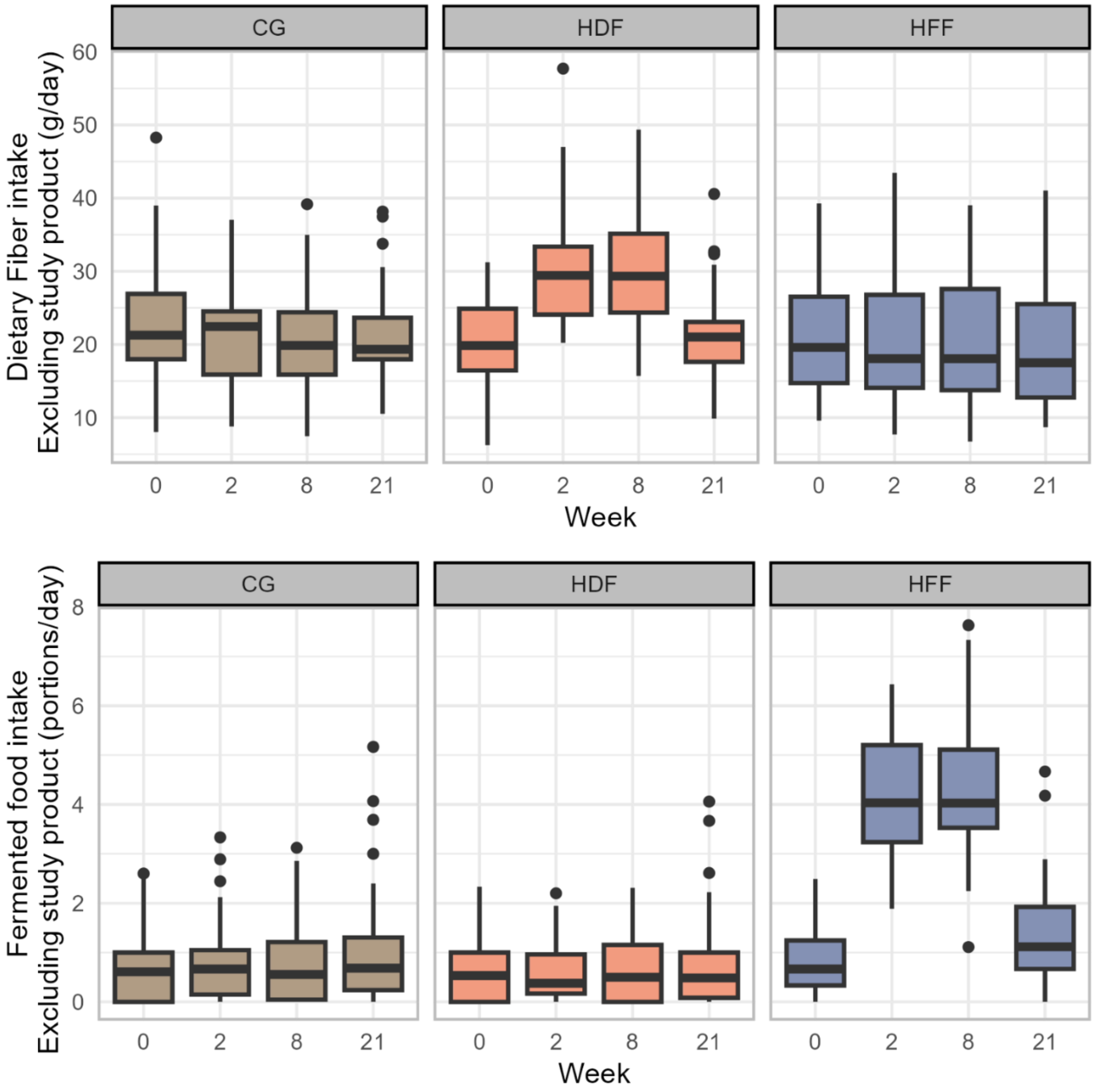


**Figure S4. Dietary fiber intake (grams/day) and fermented food intake (portions/day) excluding the intake of the study products, at baseline (week 0), end of ramp-up (week 2), end of intervention (week 8) and end of follow-up (week 21), for the PP population.** The boxplots represent the interquartile range (IQR), with the median marked by the line inside the box. Whiskers extend to the minimum and maximum scores within 1.5 times the IQR, while outliers are plotted individually with dots. Abbreviations: CG, control group; HDF, high-dietary fiber; HFF, high-fermented food; PP, per protocol.

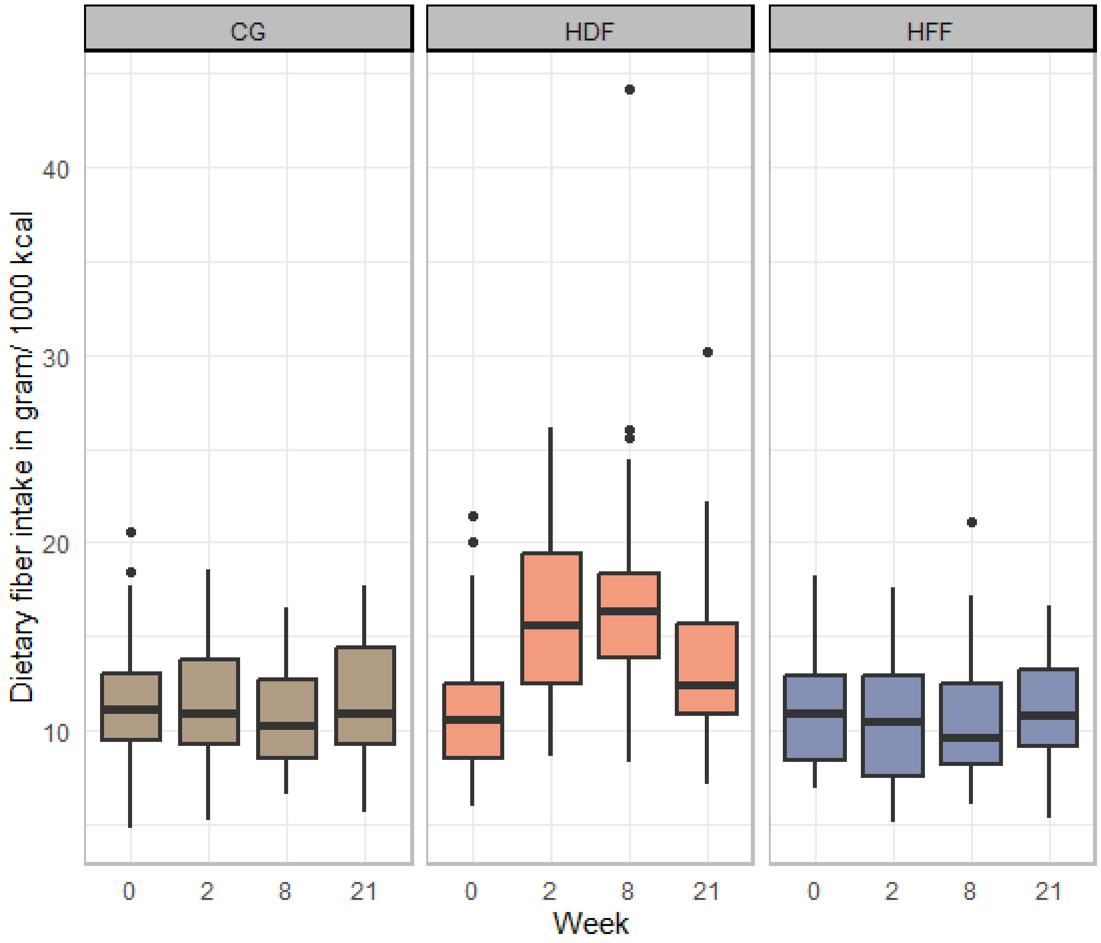


**Figure S5. Dietary fiber intake per day in gram per 1000 kilocalories, excluding the intake of the fiber study product, at baseline (week 0), end of ramp-up (week 2), end of intervention (week 8) and end of follow-up (week 21), for the PP population.** The boxplots represent the interquartile range (IQR), with the median marked by the line inside the box. Whiskers extend to the minimum and maximum scores within 1.5 times the IQR, while outliers are plotted individually with dots. Abbreviations: CG, control group; HDF, high-dietary fiber; HFF, high-fermented food; PP, per protocol.

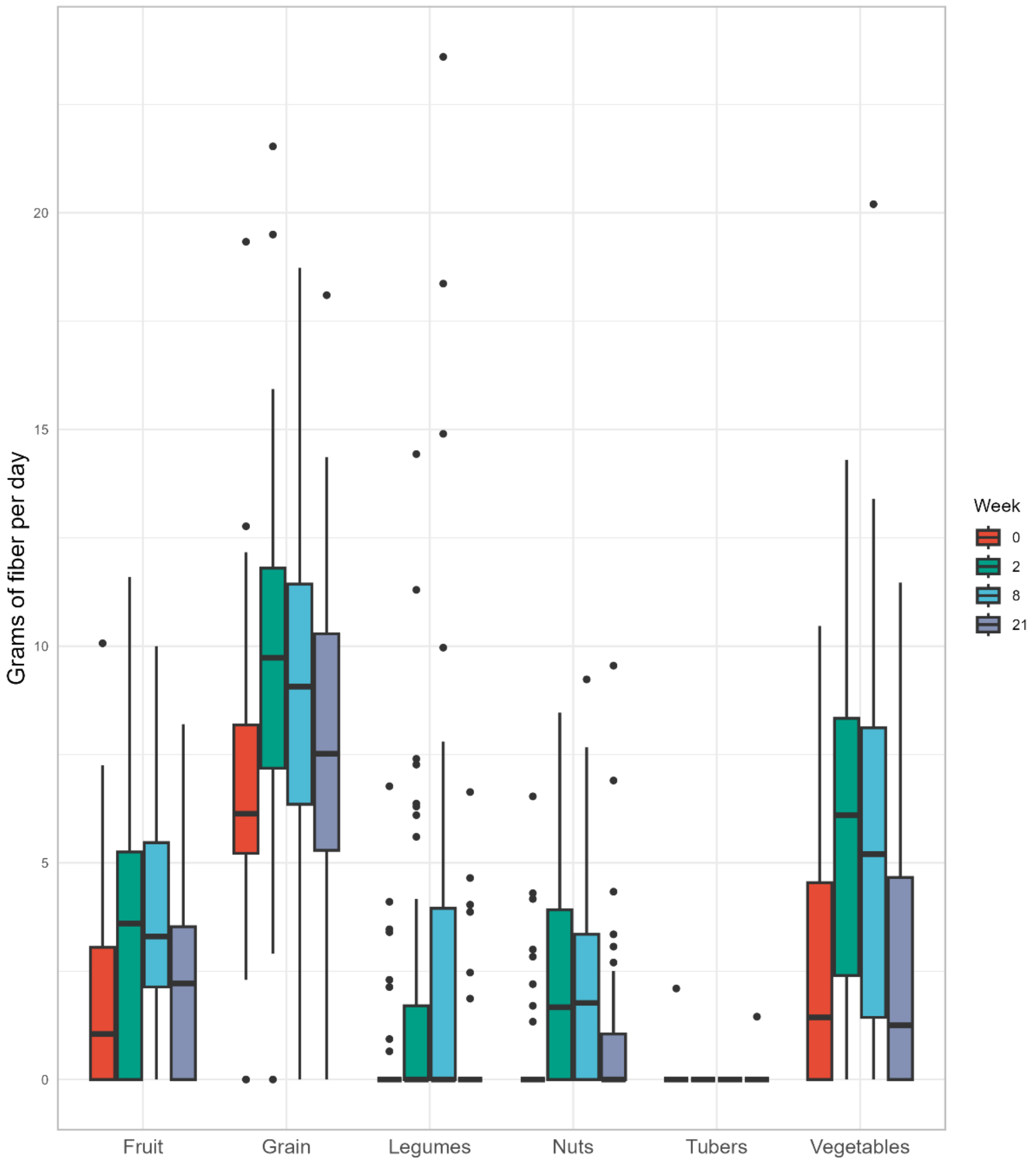


**Figure S6. Dietary fiber intake per day across categories at baseline (week 0), the end of the ramp-up phase (week 2), the end of the intervention (week 8), and the end of follow-up (week 21) in the HDF group for the PP population.** The boxplots represent the interquartile range (IQR), with the median marked by the line inside the box. Whiskers extend to the minimum and maximum scores within 1.5 times the IQR, while outliers are plotted individually with dots.Abbreviations: HDF, high-dietary fiber; PP, per protocol.

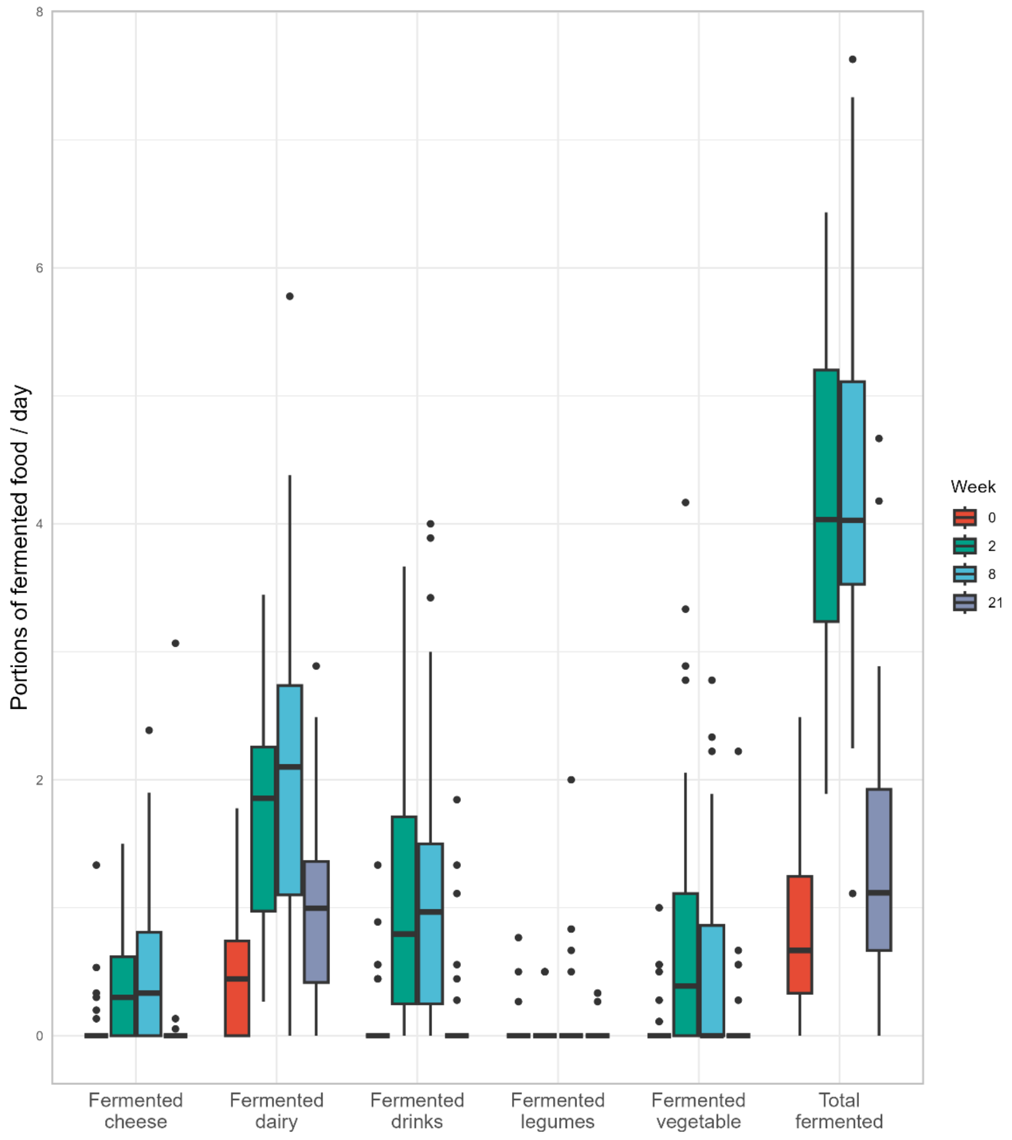


**Figure S7. Daily fermented food intake across categories at baseline (week 0), end of ramp-up (week 2), end of intervention (week 8) and end of follow-up (week 21) in the HFF group for the PP population.** The boxplots represent the interquartile range (IQR), with the median marked by the line inside the box. Whiskers extend to the minimum and maximum scores within 1.5 times the IQR, while outliers are plotted individually with dots. Abbreviations: HFF, high-fermented food; PP, per protocol.

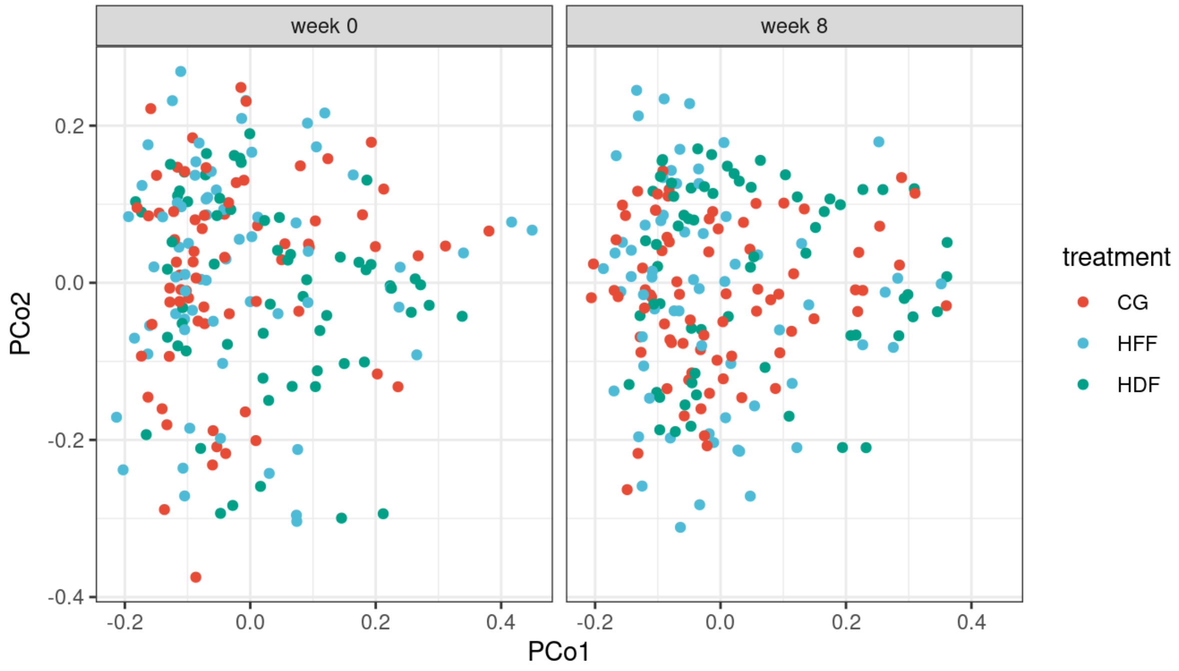


**Figure S8. Bray–Curtis β-diversity, reflected by mean distance of samples in principal coordinate plots displaying samples from week 0 and week 8, split by treatment.**

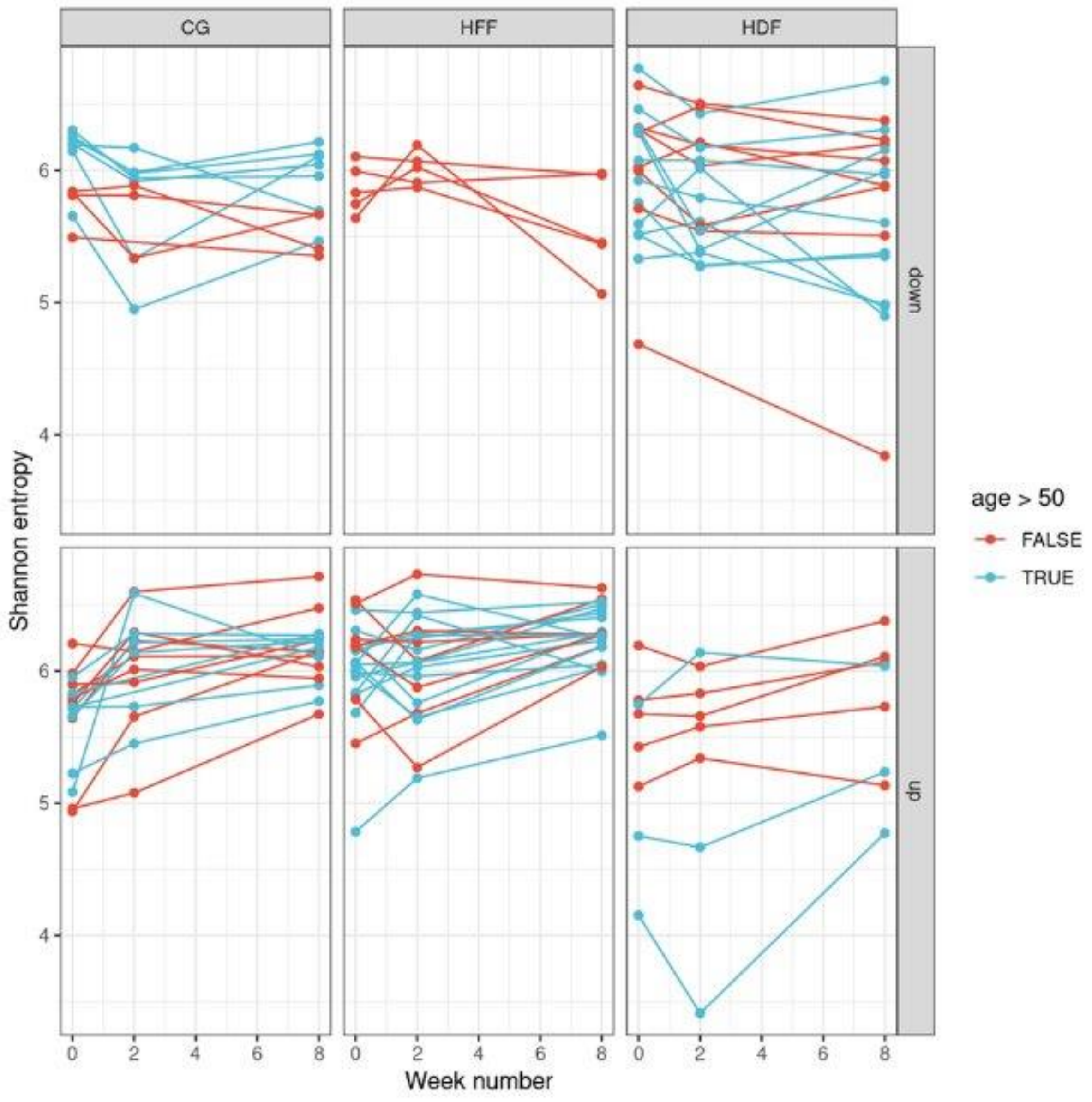


**Figure S9. Effect of age on the increase in Shannon entropy in the HFF group.** Shannon entropy going down from week0 to week8 (upper panel), or going down (lower panel). On average, participants over 50 years (blue) exhibited a more pronounced increase in Shannon diversity ($\beta$ = 0.028, *p* = 0.04) compared to those under 50 years old (in red). Abbreviations: CG, control group; HDF, high-dietary fiber; HFF, high-fermented food

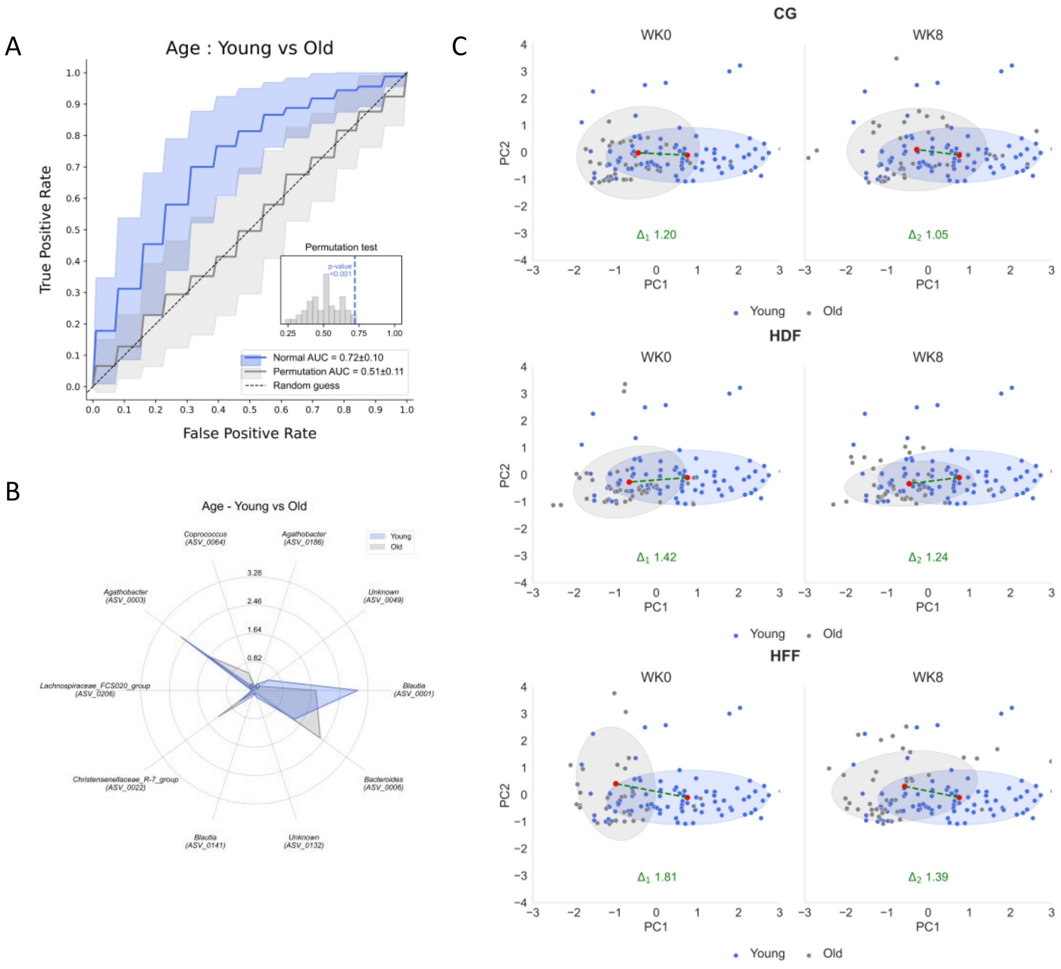


**Figure S10. Microbial age markers at baseline and shifts towards profiles associated with younger age (≤50 years old). A)** Evaluation of model performance of microbial markers associated with age at baseline by use of receiver operating characteristic (ROC) analysis, yielding an area under the curve (AUC) of 0.72 ± 0.10. To assess the robustness of this result, a permutation test was performed, generating a null distribution of AUC values (mean AUC = 0.51 ± 0.11). The observed model performance was significantly better than expected by chance (permutation *p*-value < 0.001). **B)** Radar plots presenting how abundance of the top 10 bacterial genera or species differed among study participants (*n* = 147) of > 50 years vs ≤50 years old at baseline. **C)** PCA visualization showing the microbiota composition differences between individuals younger and older than 50 years, with the centroids representing the delta shift from week 0 to week 8

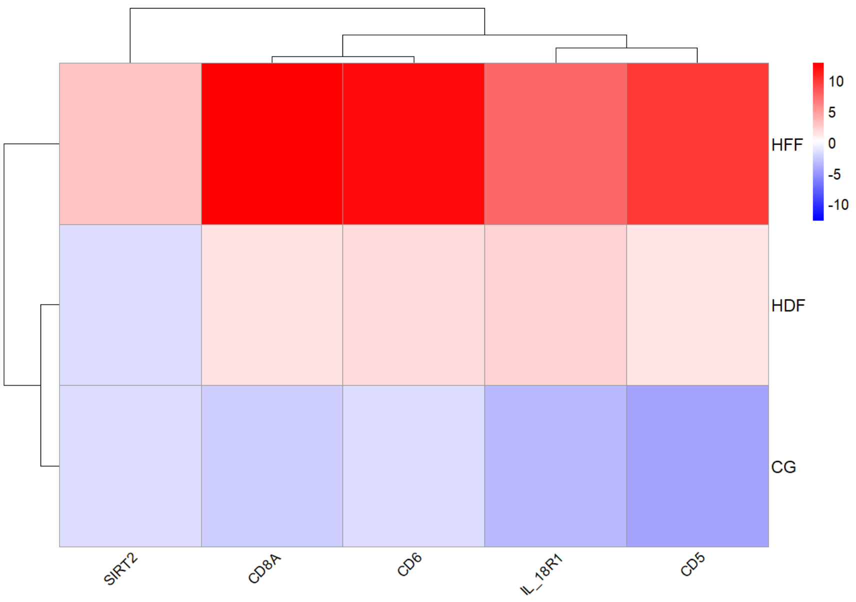


**Figure S11. Heatmap showing delta values (week8 minus week 0) for the 5 significant immune markers across HDF and HFF groups.** Hierarchical clustering was applied to the rows and columns to highlight patterns of differential abundance. Abbreviations: CG, control group; HDF, high-dietary fiber; HFF, high-fermented foods.

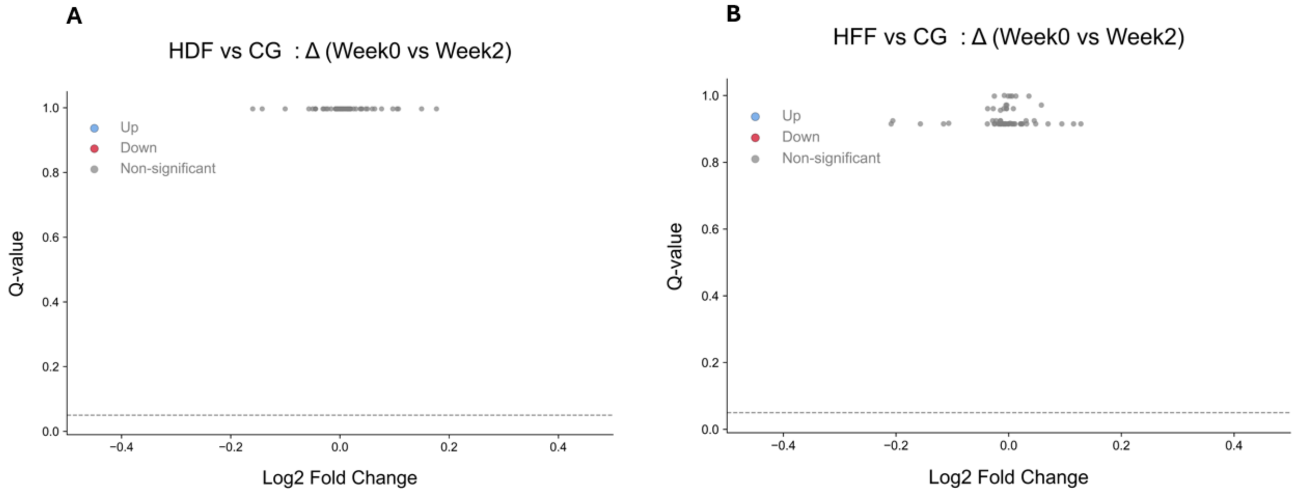


**Figure S12. Volcano plots of Log2 fold change of blood inflammation markers of HDF versus CG (A) and HFF versus CG (B) between week 2 and week 0.**

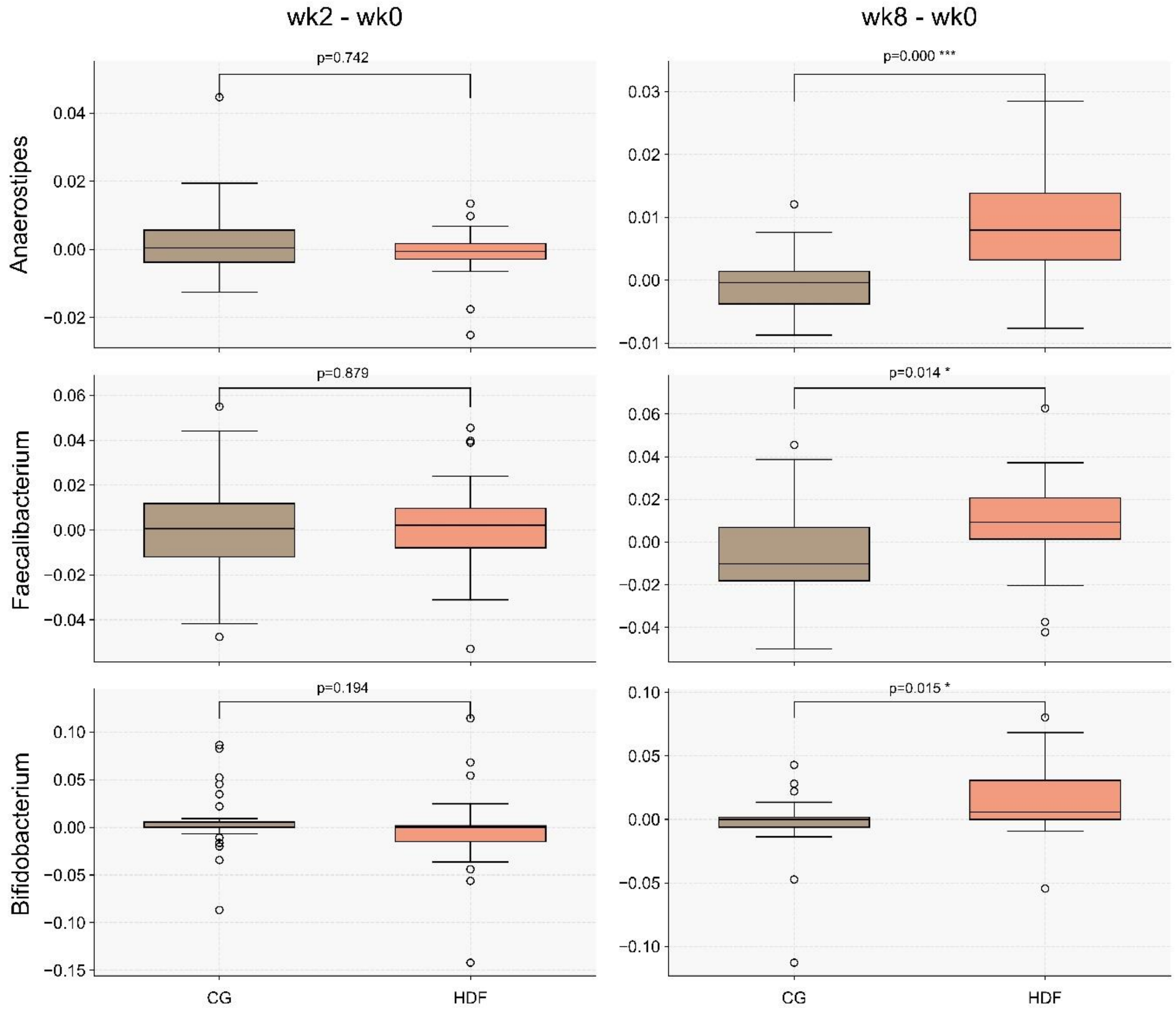

**Figure S13. Boxplots of delta values between Week 0 and Week 2 and Week 0 and Week 8 of 3 most discriminative genera between HDF and CG with a Q-value <0.05 on the Mann–Whitney U test**
